# Status of Soil-Transmitted Helminthiasis Among Adolescents in Anaocha Local Government Area, Anambra State, Nigeria: Prevalence, Associated Factors, and Future Directions After a Decade of Ongoing Mass Administration of Medicines

**DOI:** 10.1101/2023.09.15.23295620

**Authors:** Ogechukwu B. Aribodor, Eunice C. Jacob, Nwadiuto O. Azugo, Uche C. Ngenegbo, Ifeanyi Obika, Emmanuel M. Obikwelu, Obiageli J. Nebe

**Affiliations:** Department of Zoology, Nnamdi Azikiwe University, Awka, Nigeria; Social Innovation in Health Initiative (SIHI) Hub, Nigeria; Department of Parasitology and Entomology, Nnamdi Azikiwe University, Awka, Nigeria; Neglected Tropical Diseases Unit, Anambra State Ministry of Health, Awka, Nigeria; Neglected Tropical Diseases Division, Federal Ministry of Health, Nigeria

## Abstract

Over the past decade, Anambra State, Nigeria, has implemented mass administration of medicines (MAMs) to combat soil-transmitted helminthiasis (STH), a significant public health challenge in low-income regions. Nevertheless, these efforts have predominantly focused on pre-school and school-aged children, leaving a notable gap in understanding STH infection rates and the efficacy of these campaigns among secondary school adolescents, who have been excluded from this initiative. Our study aimed to address this critical knowledge gap by assessing soil-transmitted helminthiasis (STH) prevalence and contextual factors hindering effective control among adolescents in Anambra State, Nigeria. We actively engaged 443 adolescents with a mean age of 14 years in a school-based cross-sectional study in selected communities within the Anaocha Local Government Area from 8 February to 7 July 2023 following informed consent and assent procedures. Employing a stratified random sampling technique, we collected demographic data and assessed STH risk factors using a structured questionnaire hosted on the Kobo Toolbox platform. For quantitative analysis of STH infections, the Kato-Katz technique was used. Analysis was performed using SPSS version 25, incorporating descriptive statistics and multinomial logistic regression, with statistical significance set at p<0.05. Of the 443 (213 males (48.0%) and 230 females (52.0%) adolescents studied, the overall prevalence of STH observed was 35.2% (156/443). *Ascaris lumbricoides* was the prevalent STH species (16.9%), followed by *Trichuris trichiura* (1.4%) and hookworm (0.5%). Only light-intensity infection was observed. Mixed infections were observed in 16.5% of adolescents, involving *A. lumbricoides* and hookworm (10.8%), followed by *A. lumbricoides* and *T. trichiura* (3.2%) and all three STH (2.5%). The observed overall prevalence was not statistically significant with respect to gender (OR: 0.961; 95% CI: 0.651-1.420; p > 0.05) or age (OR: 0.686; 95% CI: 0.459-1.025; p>0.05). Class (grade level) (OR = 1.75, 95% CI: 1.25 - 2.45, p = 0.003), knowledge and transmission of STH infection (OR = 0.60, 95% CI: 0.42 - 0.86, p = 0.008), parental occupation (OR = 1.90, 95% CI: 1.35 - 2.67, p < 0.001), parents’ literacy level (OR = 0.68, 95% CI: 0.48 - 0.96, p = 0.027), and the type of toilet (OR = 2.15, 95% CI: 1.54 - 3.00, p < 0.001) were all significantly correlated with STH infection. These findings highlight the role of adolescents in sustaining soil-transmitted helminthiasis (STH) transmission. Coupled with school-based deworming expansion, innovative improvements in water, sanitation, hygiene, and awareness can provide a cost-effective, sustainable solution for combatting STH infections in Anambra State.

## Introduction

Soil-transmitted helminthiasis (STH) remains a persistent global health challenge, imposing a disproportionate burden on impoverished communities with limited access to sanitation and clean water [1]. These neglected tropical diseases (NTDs) are predominantly driven by three major STH species: *Ascaris lumbricoides* (roundworm), *Trichuris trichiura* (whipworm), and the hookworms, *Ancylostoma duodenale* and *Necator americanus* [2]. This group of parasitic infections collectively contributes to a staggering 85% of the global NTD burden, affecting nearly a quarter of the world’s population and resulting in 1.9 million years lived with disability (DALYs) [3–4].

Regions characterized by poor environmental sanitation, inadequate personal hygiene, limited educational opportunities, low socio-economic status, and constrained access to healthcare and safe drinking water consistently report higher STH prevalence rates [5]. Additionally, children born to mothers infected with these helminths during pregnancy are at elevated risk of infection during their early years [6]. Practices such as geophagy, open defecation, the use of leaves and paper for hygiene, nail-biting, and walking barefoot further contribute to the persistently high prevalence of STH infections [7–9].

Sub-Saharan African nations, notably Nigeria, bear the brunt of STH infections, with significant disparities across regions. Notably, southern regions grapple with higher prevalence rates compared to their northern counterparts [10].

The cornerstone of STH control lies in mass drug administration (MDA), complemented by health and hygiene education, and the provision of clean water and sanitary facilities to ensure the sustained impact of MDA. However, the effectiveness of these measures is hampered by the oversight of specific demographic groups, particularly adolescents, who often escape targeted interventions, leading to reinfection among treated children and others. In the African context, adolescents frequently assume caregiving roles in the absence of parents and play a pivotal role in shaping the health and well-being of younger individuals. Thus, their adoption of healthy practices and their role in educating others is of paramount significance.

While numerous studies have examined STH infections in Anambra State, a conspicuous gap remains, specifically regarding the burden of STH among adolescents [11–14]. Against the backdrop of a decade-long MDA initiative in Anambra State, it is imperative to furnish evidence-based insights into the STH status of adolescents, who represent the future workforce of both the state and Nigeria. Their health and well-being are intricately linked to the nation’s economic prosperity. Hence, this study diligently evaluates the endemicity of soil-transmitted helminthiasis and identifies the multifaceted factors that impede its effective control among adolescents in Anambra State.

## Materials and Methods

### Study area

The study was carried out in the Anaocha Local Government Area (L.G.A.), situated in Anambra State, within the southeastern region of Nigeria (Fig. 1). Anaocha L.G.A. is one of the twenty-one Local Government Areas in the state, spanning an area of approximately 171.62 square kilometres and hosting an estimated population of 418,360 distributed across ten communities [15–16]. Two communities were randomly selected from the LGA namely, Adazi-Nnukwu and Agulu by ballot method to mitigate bias [17].

**Fig. 1:**
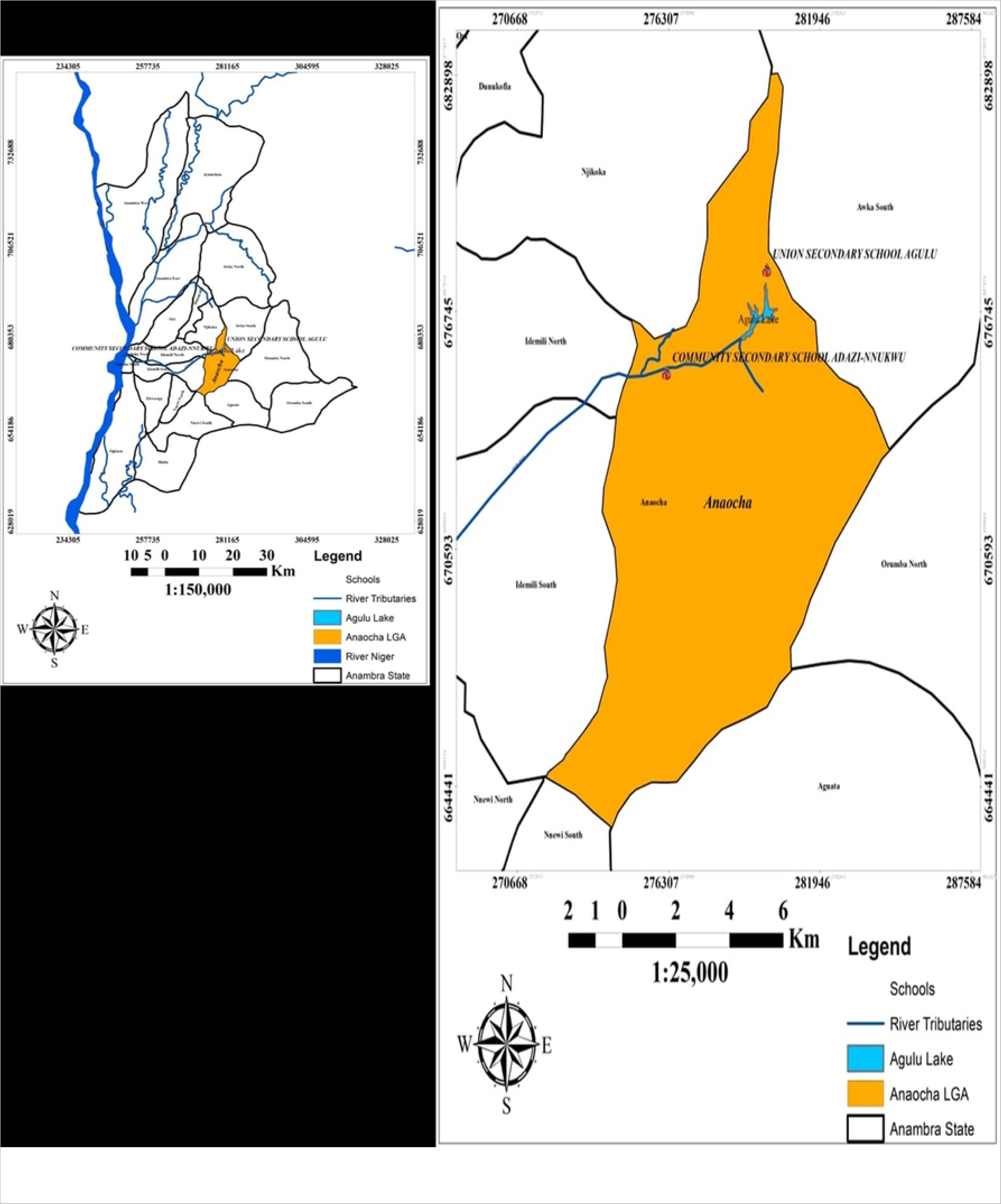
Map of the Anambra State Showing the Selected Local Government Area and the Locations Chosen for the Study (Source: Nnamdi Azikiwe University, Geography Information System Laboratory, 2023)

Adazi-Nnukwu, positioned at coordinates 6.1018° N and 7.0122° E within Anaocha LGA, is a community nestled within the Nri-Awka Area. In contrast, Agulu, located at coordinates 6.1160° N and 7.0678° E, serves as the largest town within Anaocha LGA. The populace is predominantly of the Igbo ethnic group who speak both Igbo and English languages. Christianity and traditional religions are the main religions practised in this area. The study area’s vegetation is classified under the tropical rainforest zone, though it has been significantly altered by various human activities, resulting in a remnant of trees and grasses. The region experiences a humid climate characterized by two distinct seasons: the dry season spanning from November to March and the rainy season from April to October. Topographic features in the study area encompass uplands, lowlands, stream channels, and gullies of variable dimensions. The elevation above sea level in both communities ranges from 65 metres to 212 metres. The average temperature is estimated at 25.5°C, and an annual rainfall of approximately 1,897 millimetres with relative humidity peaking at 85% during the rainy season and averaging 65% during the dry season. The majority of residents are farmers, traders, civil servants and teachers. The health centres and hospitals present in the L.G.A. grapple with critical challenges, chief among them being the shortage of qualified healthcare personnel and the absence of well-equipped laboratories. Also, there is a need for improved sanitary infrastructure as open defecation is a prevalent practice in these communities due to the lack of accessible public toilets. Additionally, it is pertinent to note that a significant majority of government-owned schools, commonly referred to as public schools, are often perceived as institutions catering primarily to economically disadvantaged students [13], further accentuating the complex issues surrounding access to quality education and healthcare within these areas.

### Study design

A school-based, cross-sectional study was conducted from 8 February to 7 July 2023. Only government-owned secondary schools that had a mixed population of male and female students were enrolled in the study. Eligible participants were adolescents aged 10 to 19 years who had resided in the chosen communities for at least three years and were enrolled in the selected secondary schools. Adolescents who were on medications or had taken anthelminthic drugs within a month before the commencement of the study were excluded from this study.

### Study population

The study population consisted of secondary school students from junior secondary classes 1-3 and senior secondary classes 1-2 enrolled in government-owned secondary schools namely: Union Secondary School, Agulu, and Community Secondary School, Adazi-Nnukwu who were aged between 10-19 years and had provided consent to participate in the study. The senior secondary class 3 was not enrolled in the study due to the ongoing West African Senior School Certificate Examination (WASSCE). This exclusion was necessary as the scheduled exam times conflicted with the time for collecting samples.

### Sample size and sampling technique

The sample size was determined using the single population proportion formula for health studies [18] and assuming a 44.2% prevalence rate of STH derived from a previous study conducted in Anambra State [12].

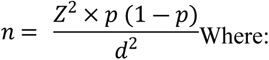

n= minimum sample size

Z= reliability coefficient= 1.96 at 95% confidence interval

p= Expected prevalence probability of success = 44.2% = 0.442.

d = margin of error = 5% = 0.05

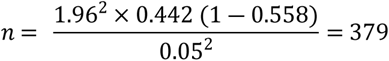

The minimum sample size required for this study was 379. However, the sample size was increased to 443 to maximize precision and mitigate errors from possible withdrawals and missed data.

Using the stratified random sampling technique [19], participants were randomly selected from 5 strata based on their classes, ranging from Junior Secondary School (JSS) to Senior Secondary School (SSS) (JSS 1 – SSS 2).

### Data collection

Oral interviews and a pre-tested questionnaire were used to collect information on socio-demographic characteristics and risk factors for STH. The questionnaire data was collected using the Kobo toolbox during the same period as the stool samples, between 8:00 a.m. and 12:00 p.m. Information was collected on age, class, parents’ occupation, parents’ literacy level, availability of toilet at home and the type of toilet available, source of drinking water, availability of water facilities and hand washing facilities in the school and home, and hygiene behaviour. Knowledge about STH was documented with emphasis on (i) awareness, (ii) source of information, and (iii) transmission routes. We also documented (i) if adolescents have ever been dewormed, (ii) when they were last dewormed, and (iii) who administered the medicine. In addition to this, focus group discussions (FGDs) were employed to gather more insights, where the adolescents were divided into groups of five based on gender and age.

### Stool sample collection and examination

Stool samples were collected from each adolescent into labelled, sterilized, dry, leak-proof, and screw-cap stool containers. To ensure proper identification, each container was accurately labelled and coded according to the participant’s identity. The collected specimens were checked to ensure the correctness of labels, quantity, and collection procedure. Stool sample collection was done between 8:00 a.m. and 12:00 p.m. to ensure that the samples were fresh and suitable for analysis. The stool samples were then stored in an ice-packed box and transported promptly to the Laboratory of the Department of Zoology, Nnamdi Azikiwe University, Awka, Anambra State, Nigeria within a maximum of an hour to prevent degradation and contamination. The standard Kato-Katz technique was used to examine stool specimens for the presence of *A. lumbricoides*, *T. trichiura*, and hookworm eggs. The prepared slides were examined under the microscope with ×10 and × 40 magnifications. All eggs detected in the preparations were identified, counted, and multiplied by a factor of 24, with the results represented as eggs per gram of stool (EPG) for the intensity of infection, according to WHO criteria [20]. All the Kato-Katz slides were prepared immediately after the stool specimens were received in the laboratory and read in line with the WHO-recommended reading time (30 to 60 minutes).

### Quality Control/Assurance

The completeness of the questionnaire and the quantity of the sample were checked daily. Data collectors were supervised during data collection. For egg quantification, each prepared slide was examined by a laboratory technologist and the researcher, and the average result was taken. Stool samples were randomly selected for quality control and re-examined by a laboratory technologist and the results were recorded separately and compared with the original egg count data.

### Data analysis

The collected data from the questionnaires were checked for completeness and consistency using Kobo toolbox. The data was then exported in Excel format from Kobo toolbox to Microsoft Excel, where it was coded, and imported into the Statistical Package for the Social Sciences software version 25 (SPSS) for statistical analysis. Descriptive statistics such as frequencies and percentages were used to describe the demographic characteristics of the study participants. The mean intensity of the parasite EPG of stool was calculated for all infected and non-infected individuals. Multinomial logistic regression analyses were employed to identify the association of each independent variable with the dependent variable. The strength of the association between risk factors and STH infection was tested by odds ratio with 95% confidence intervals. In all cases, a p-value less than 0.05 was considered statistically significant.

### Ethical approval and informed consent

Ethical clearance was obtained from the Health Research and Ethics Committee of Chukwuemeka Odumegwu Ojukwu University Teaching Hospital (COOUTH), Anambra (ref no: COOUTH/CMAC/ETH.C/Vol.1/FN:04/247) and an official permission was sought from Post Primary Schools Service Commission. The parents, guardians and teachers were sensitized during the Parents Teachers Association meetings in the selected schools on the aim and objectives of this research. Written informed consent was obtained from adolescents aged 18 to 19 years. Adolescents aged between 10 to 17 years gave assent to participate and completed assent forms through their parents or legal guardians.

## Results

### Socio-demographic characteristics and hygiene conditions of the adolescents studied

As presented in Table 1, a total of 443 adolescents aged 10 – 19 years old were recruited for this study from the schools visited. Among them, 51.9% (230/443) were females and 48.1% (213/443) were males. The mean age of the study participants was 14.09 years (SD: ±2.47 years). Adolescents between the ages of 10-14 years were 58.2% (258/443), while 41.8% (185/443) were between the ages of 15-19 years. Also, 61.4% (272/443) and 38.6 (171/443) of the adolescents studied were from Agulu and Adazi-Nnukwu respectively. Most of the study participants (65.9%, 292/443) had parents who were traders/entrepreneurs. A majority of them had parents who had completed secondary school education, 63.7% (282/443). There was no toilet facility in the schools.

**Table 1:**
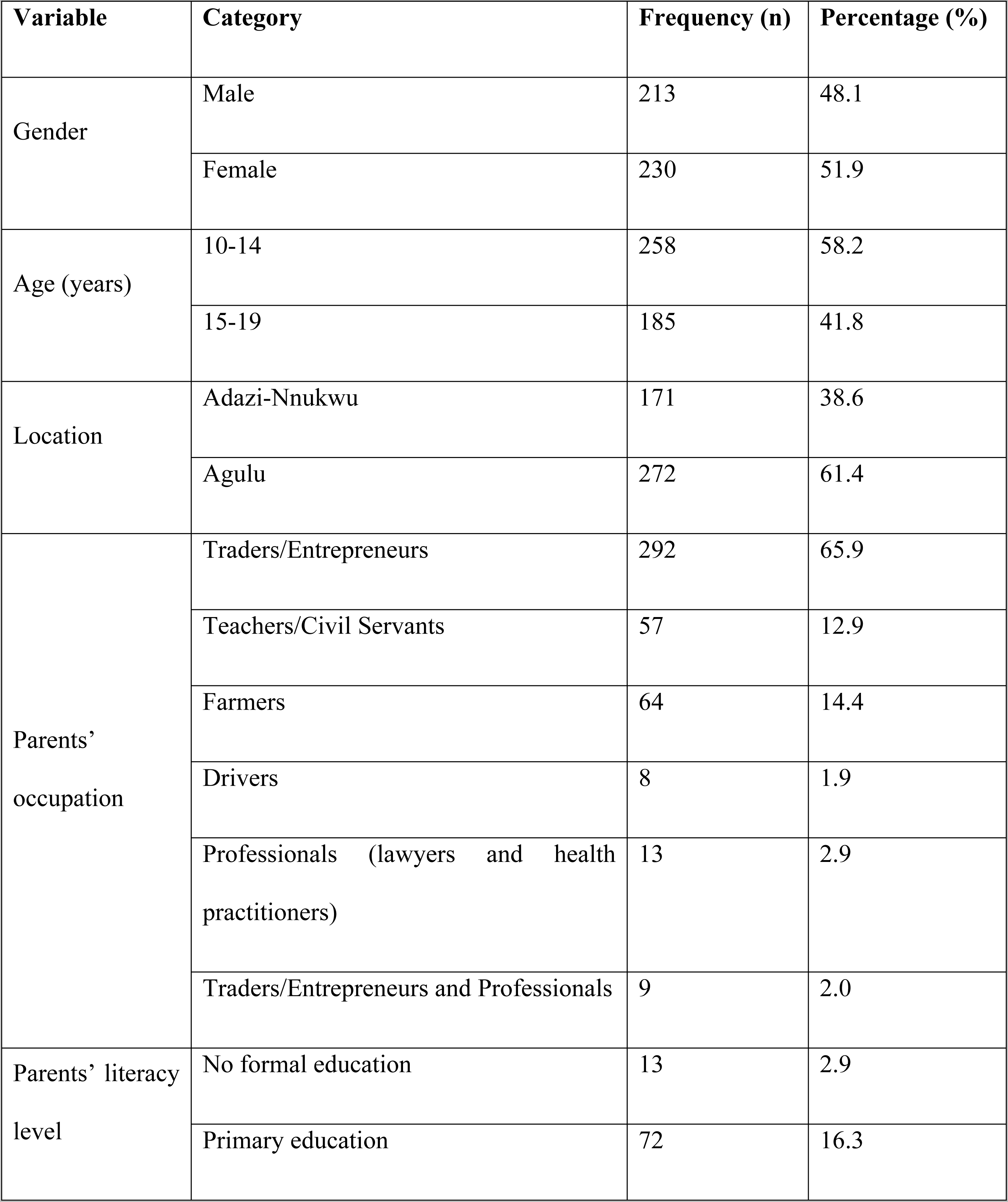
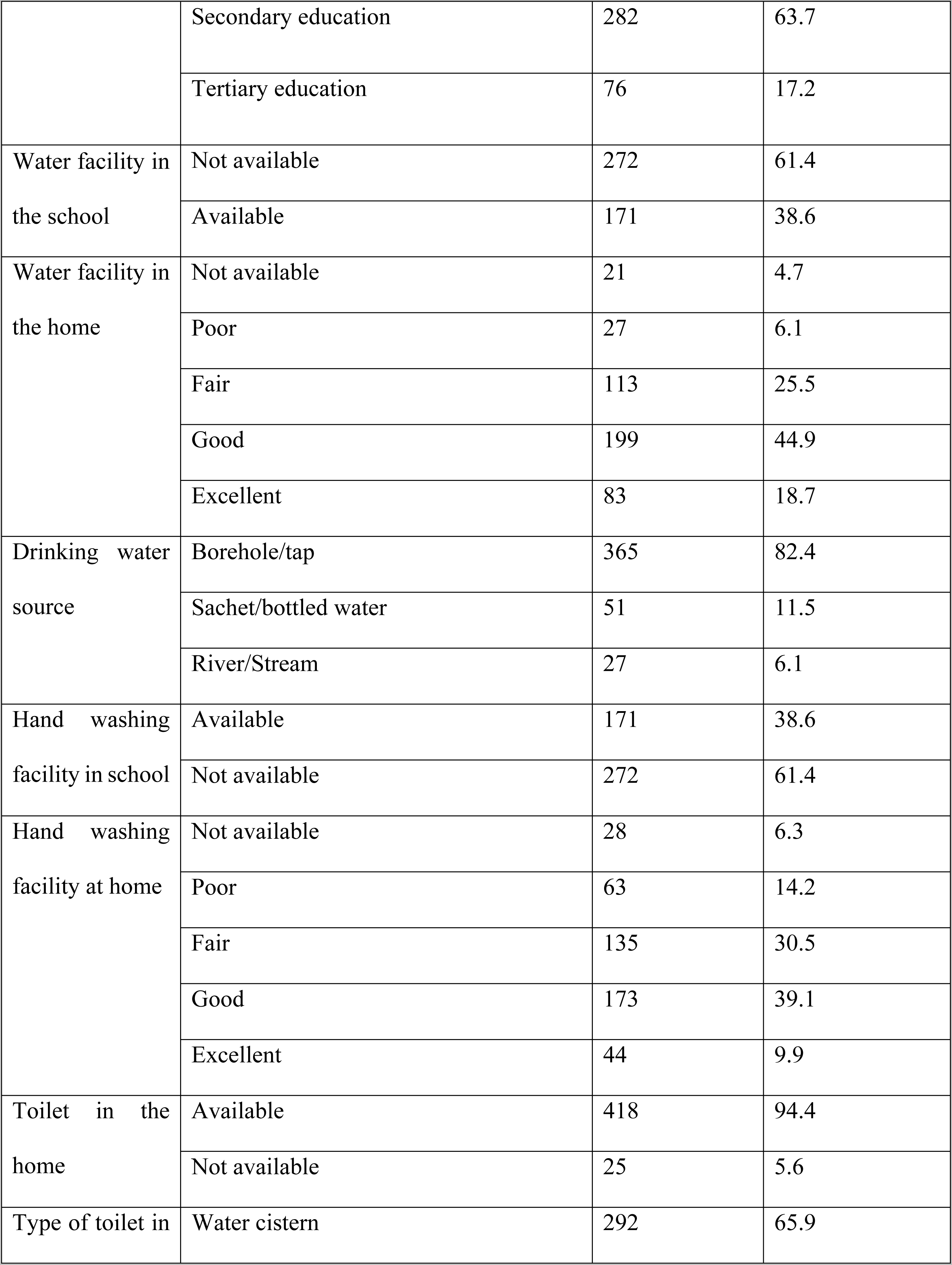
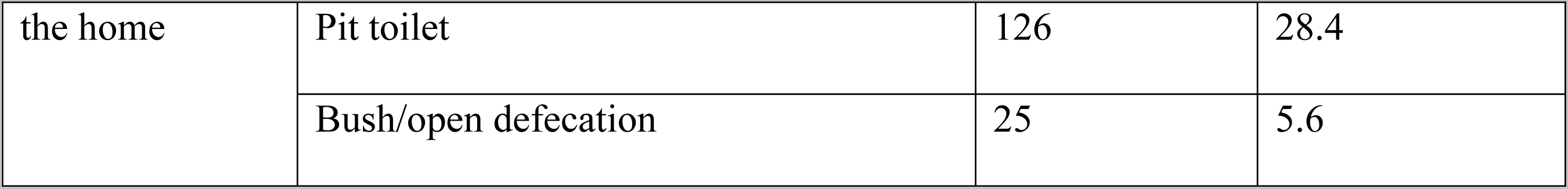
Socio-demographic characteristics of adolescents in Anaocha L.G.A., Anambra State, Nigeria (n = 443)

### Prevalence of soil-transmitted helminths infection among adolescents studied

The overall prevalence of STH infection among adolescents studied was found to be 35.2% (156/443) (95% CI: 30.6-39.7). Parasite-specific infection prevalence was 16.9% (75/443) for *A. lumbricoides*, 1.4% (6/443) for *T. trichiura*, and 0.5% (2/443) for hookworm. Most of the adolescents had a single infection 18.7% (83/443) while 16.5% (73/443) of them had multiple infections, with the most prevalent being *A. lumbricoides* and hookworm 10.8% (48/443), followed by *A. lumbricoides* and *T. trichiura* 3.2% (14/443), and *A. lumbricoides*, *T. trichiura*, and hookworm 2.5% (11/443). The prevalence of STH infection was slightly higher in males 35.7% (76/213) than in females 34.8% (80/230) (OR: 0.961; 95% CI: 0.651-1.420; p=0.843) and in adolescents aged 10-14 years 38.8% (100/258) (OR: 0.686; 95% CI: 0.459-1.025; p=0.066) (Table 2). The prevalence of STH parasites among adolescents who knew about STH infections was found at 40.3% (124/308). Of the 9.7% (43/443) who got this information from MAMs, 58.1% (25/43) were infected with STH (Fig. 2.)

**Fig. 2:**
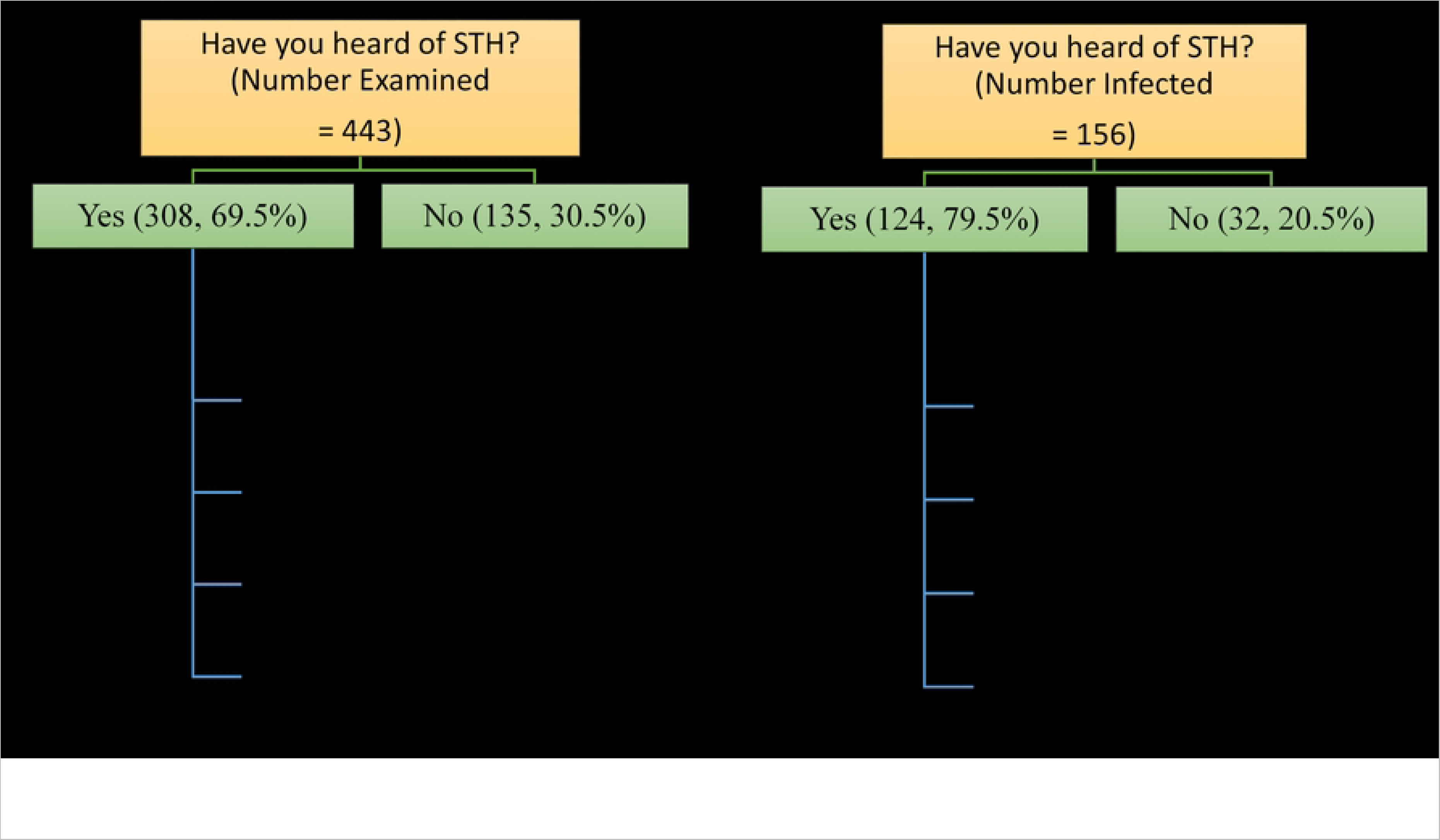
Awareness and prevalence of soil-transmitted helminthiasis among adolescents studied in Anaocha L.G.A., Anambra State.

**Table 2:**
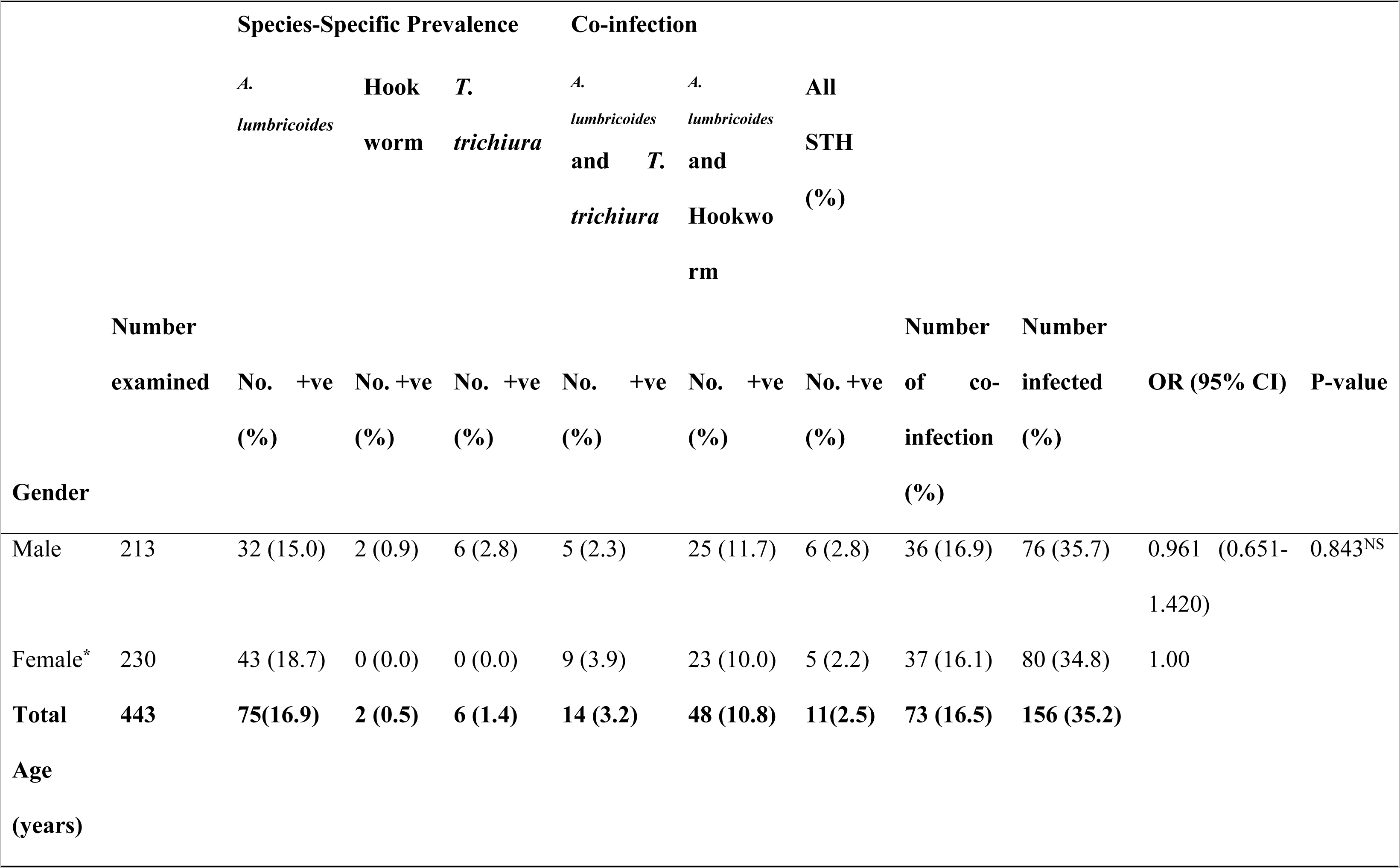
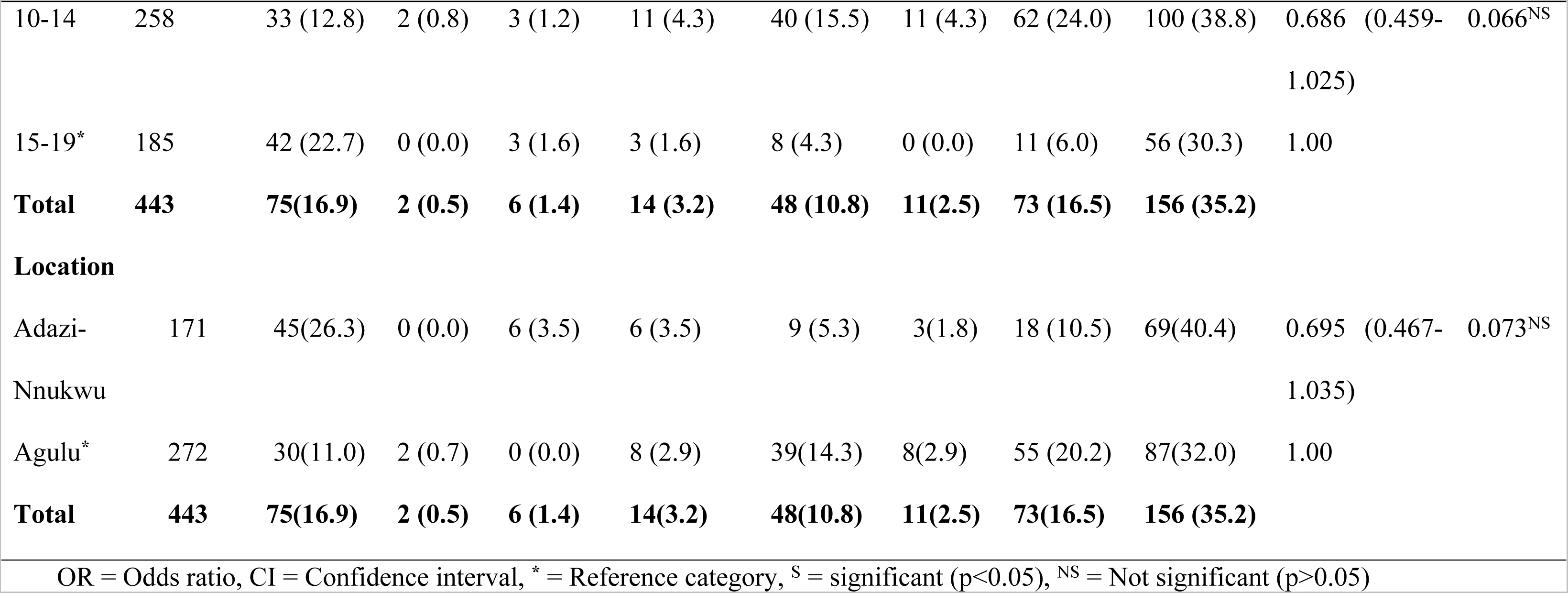
Species-specific prevalence of STH among adolescents studied in Anaocha L.G.A., Anambra State, Nigeria (n = 443)

### Intensity of soil-transmitted helminths infection among adolescents studied

Results showed that all 156 adolescents with STH infections had light-intensity infections of *A. lumbricoides*, hookworm, and *T. trichiura*. The overall mean EPG of *A. lumbricoides* was 109.14 (95% CI: 95.77-122.50). However, female adolescents had a higher arithmetic mean EPG of 109.4 (95% CI: 92.14-126.66) than males, whose mean EPG was 108.85 (95% CI: 87.77-129.92). Meanwhile, male adolescents infected with *T. trichiura* had a higher mean EPG of 39.43 (95% CI: 27.76-51.09) compared to females whose mean EPG was 32.47 (95% CI: 26.39-38.55). The overall mean EPG for *T. trichiura* was 35.61 (95% CI: 29.65-41.57), and for hookworm, it was 59.02 (95% CI: 32.05-85.99) (Table 3).

**Table 3:** Mean intensity of STH among adolescents studied in Anaocha L.G.A., Anambra State, Nigeria.

| STH | Category | Number Infected (%) | Arithmetic Mean EPG | Range | 95% Confidence Interval |
| --- | --- | --- | --- | --- | --- |
| *A. lumbricoides* | Sex |  |  |  |  |
|  | Male | 71 (48.0) | 108.85 | (24-360) | (87.77-129.92) |
|  | Female | 77 (52.0) | 109.4 | (24-360) | (92.14-126.66) |
|  | Age |  |  |  |  |
|  | 10-14 years | 98 (66.2) | 104.33 | (24-360) | (89.0-119.65) |
|  | 15-19 years | 50 (33.8) | 118.56 | (24-360) | (92.22-144.9) |
|  | Community |  |  |  |  |
|  | Adazi-Nnukwu | 63 (42.6) | 116.57 | (24-360) | (94.38-138.76) |
|  | Agulu | 85 (57.4) | 103.62 | (24-360) | (86.86-120.38) |
|  | **Overall** | **148 (100)** | **109.14** | **(24-360)** | **(95.77-122.50)** |
| *T. trichiura* | **Sex** |  |  |  |  |
|  | Male | 14 (45.2) | 39.43 | (24-72) | (27.76-51.09) |
|  | Female | 17 (54.8) | 32.47 | (24-48) | (26.39-38.55) |
|  | **Age** |  |  |  |  |
|  | 10-14 years | 22 (71.0) | 30.55 | (24-48) | (25.69-35.40) |
|  | 15-19 years | 9 (29.0) | 48.0 | (24-72) | (32.02-63.98) |
|  | **Community** |  |  |  |  |
|  | Adazi-Nnukwu | 15 (48.4) | 38.4 | (24-72) | (27.39-49.41) |
|  | Agulu | 16 (51.6) | 33.0 | (24-48) | (26.61-39.39) |
|  | **Overall** | **31 (100)** | **35.61** | **(24-72)** | **(29.65-41.57)** |
| **Hookworm** | **Sex** |  |  |  |  |
|  | Male | 33 (54.1) | 47.27 | (24-120) | (35.72-58.83) |
|  | Female | 28 (45.9) | 72.86 | (24-840) | (14.03-131.69) |
|  | **Age** |  |  |  |  |
|  | 10-14 years | 53 (86.9) | 60.68 | (24-840) | (29.61-91.75) |
|  | 15-19 years | 8 (13.1) | 48.0 | (24-72) | (29.42-66.58) |
|  | **Community** |  |  |  |  |
|  | Adazi Nnukwu | 12 (19.7) | 36.0 | (24-48) | (28.04-43.96) |
|  | Agulu | 49 (80.3) | 64.65 | (24-840) | (31.08-98.22) |
|  | **Overall** | **61 (100)** | **59.02** | **(24-840)** | **(32.05-85.99)** |

### Risk factors of soil-transmitted helminths infections among adolescents studied

The results showed that adolescents who had no knowledge about STH infection (OR: 4.286; 95% CI: 1.423-12.906; p<0.05) were significantly 4.3.times more likely to be infected with one or more species of STH than those who had prior knowledge (Fig. 3) (Table 4). Furthermore, adolescents who received education about STH infections in school (OR: 0.455; 95% CI: 0.276-0.749; p<0.05) and through MAMs (OR: 0.224; 95% CI: 0.108-0.461; p<0.05) were less likely to be infected compared to those who had no knowledge of the infection (Fig. 3) (Table 4).

**Fig. 3:**
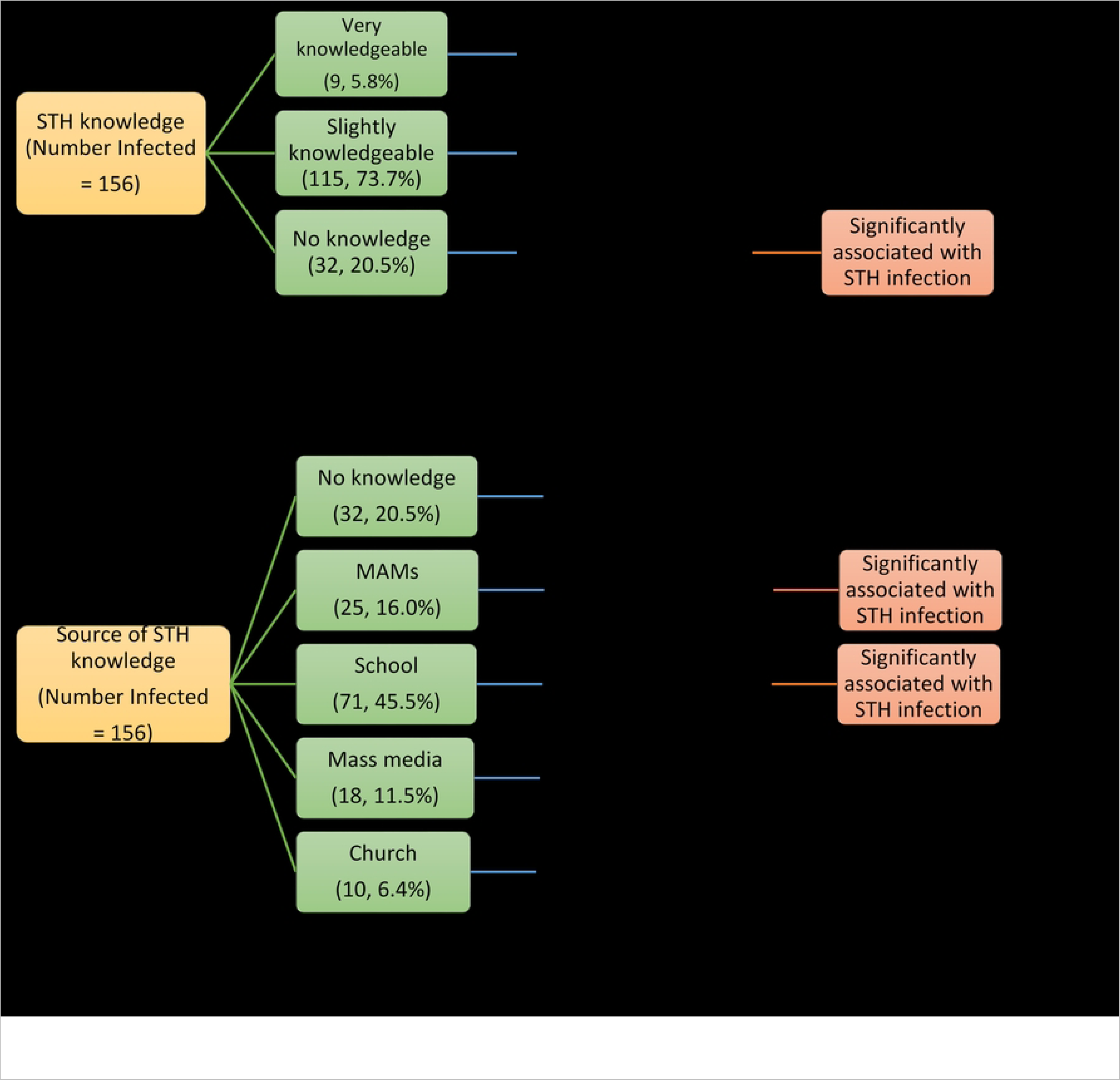
Association of knowledge and source of knowledge about soil-transmitted helminths with soil-transmitted helminths infection among adolescents studied in Anaocha L.G.A., Anambra State.

**Table 4:**
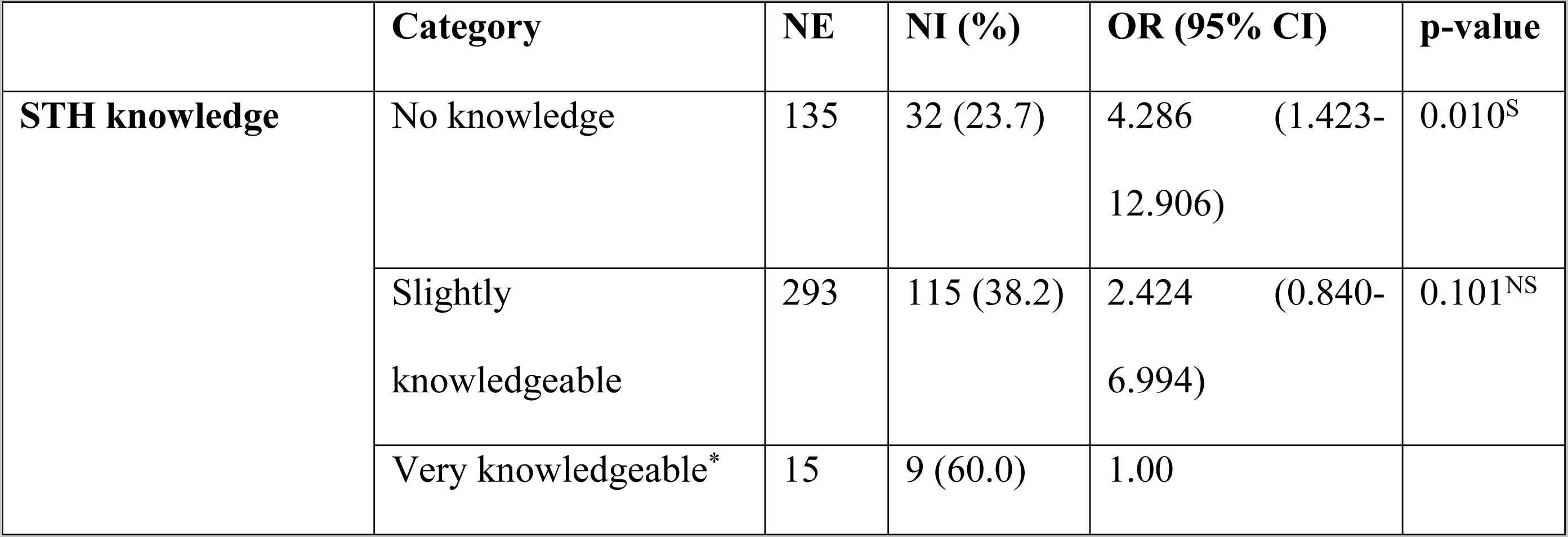
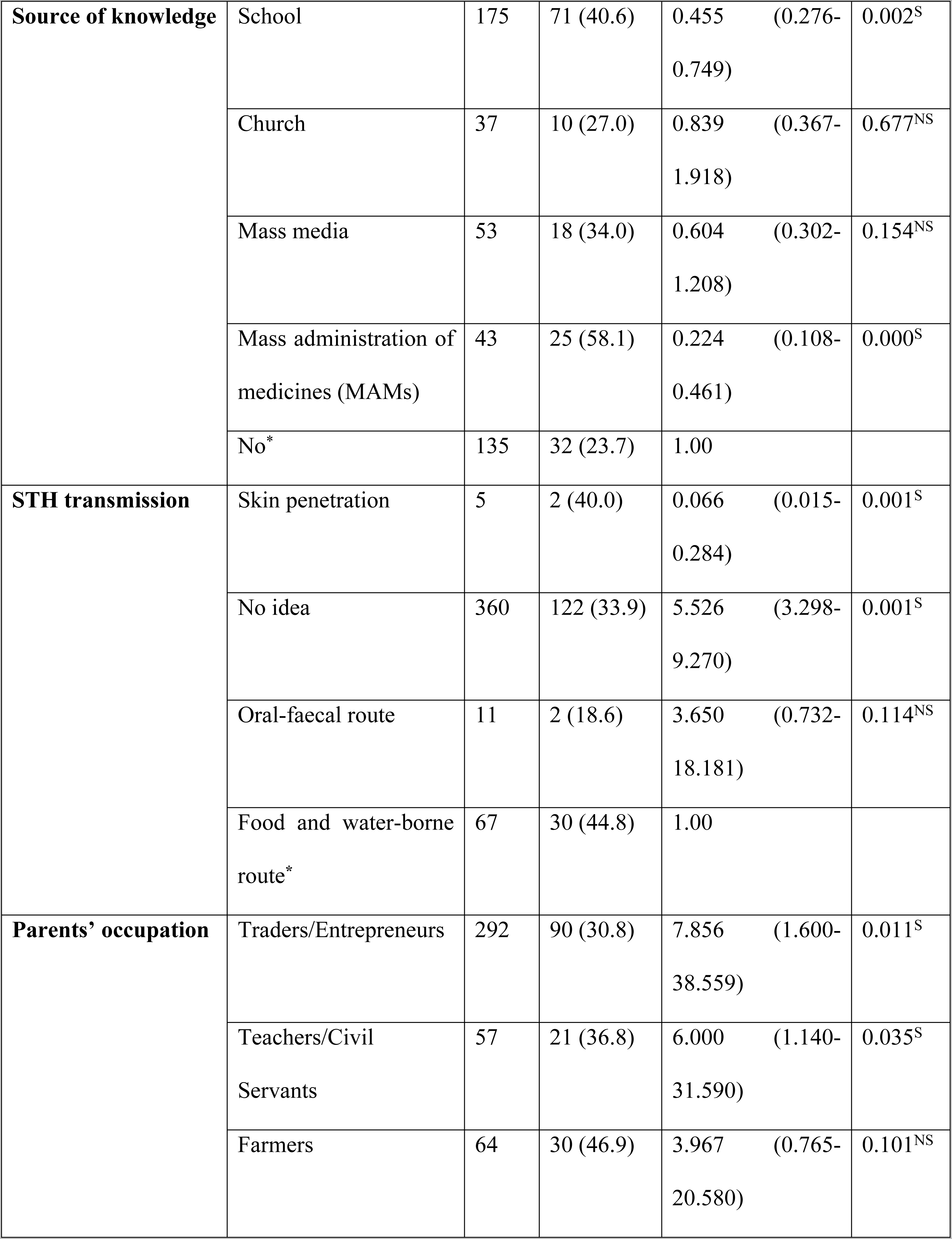
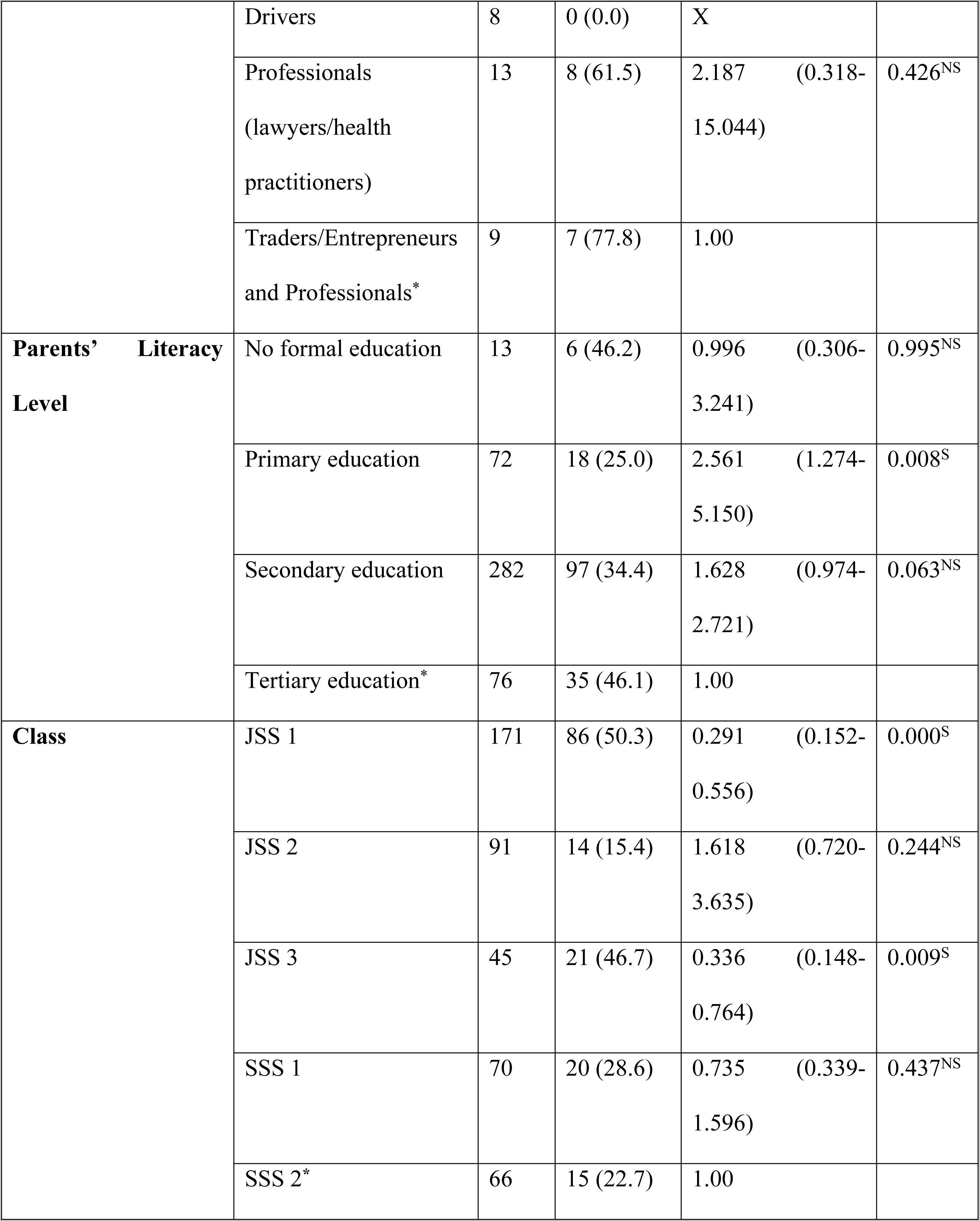
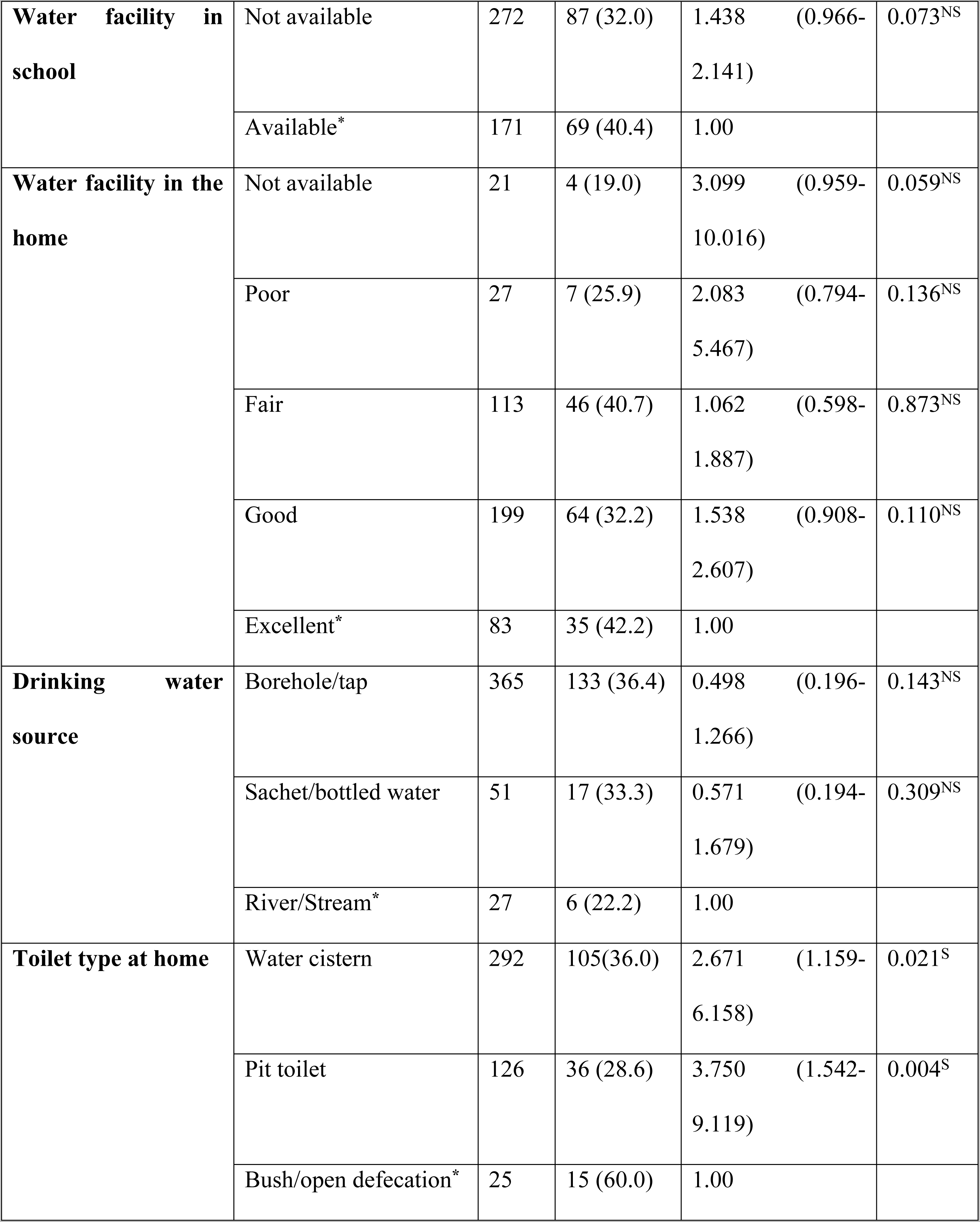
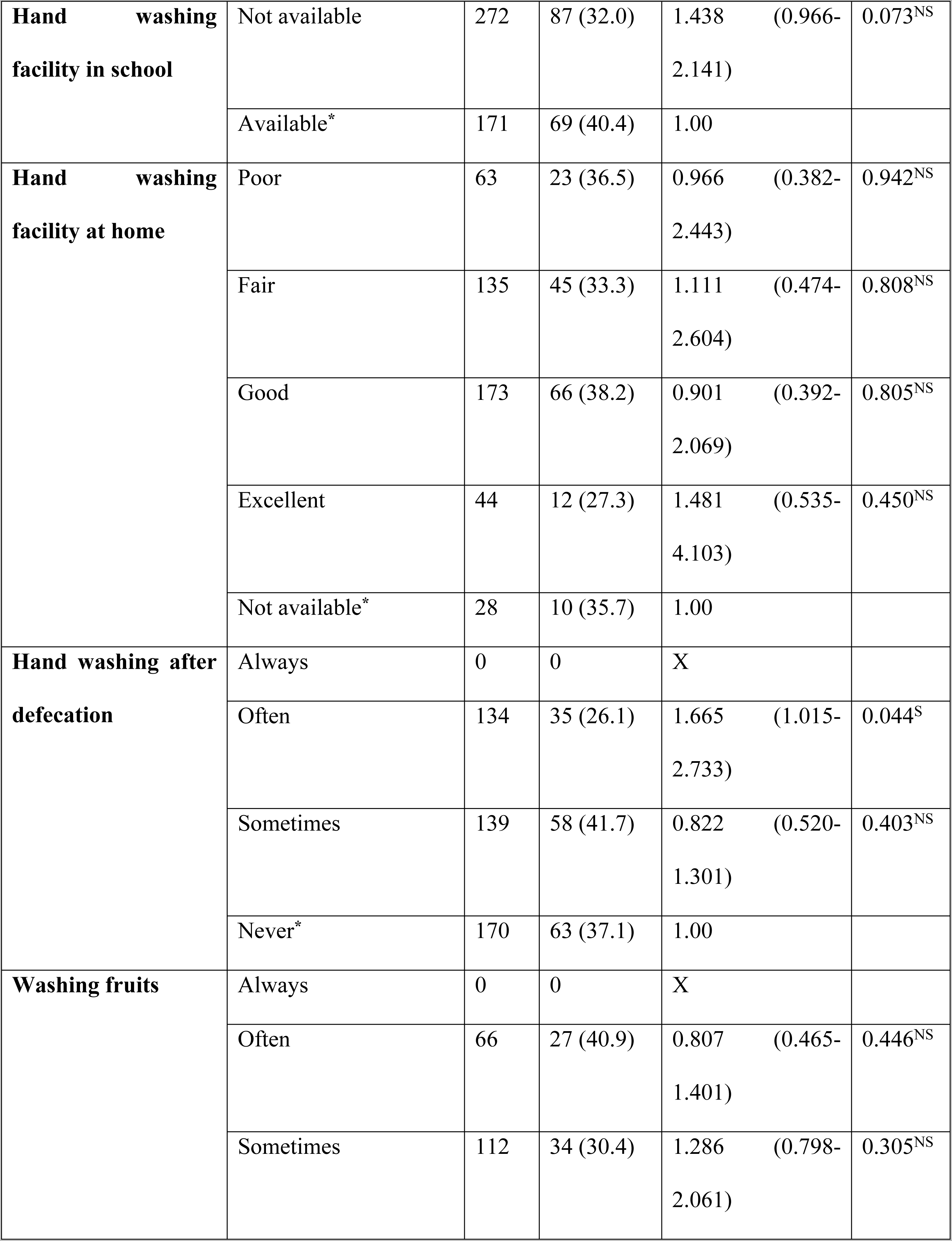
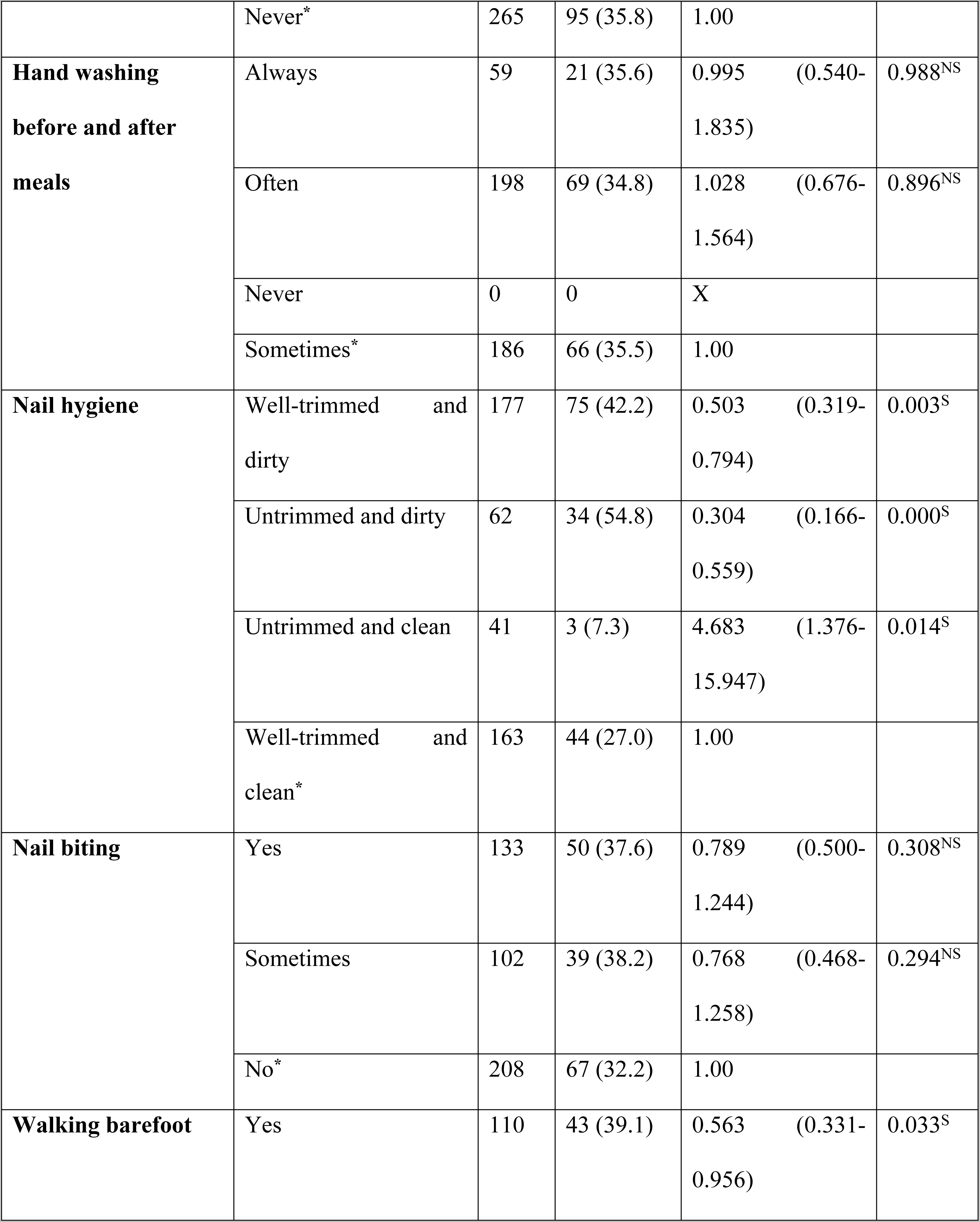
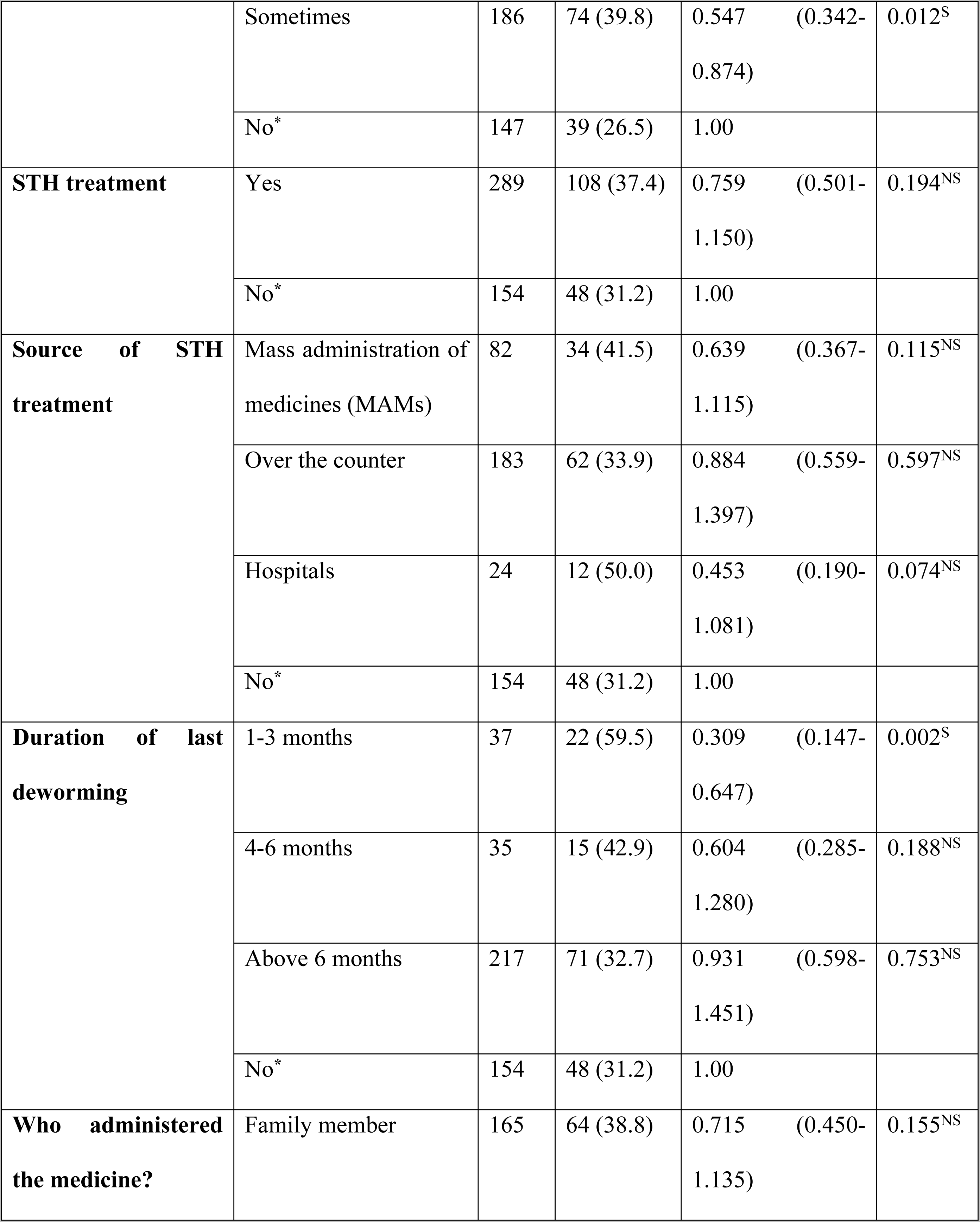
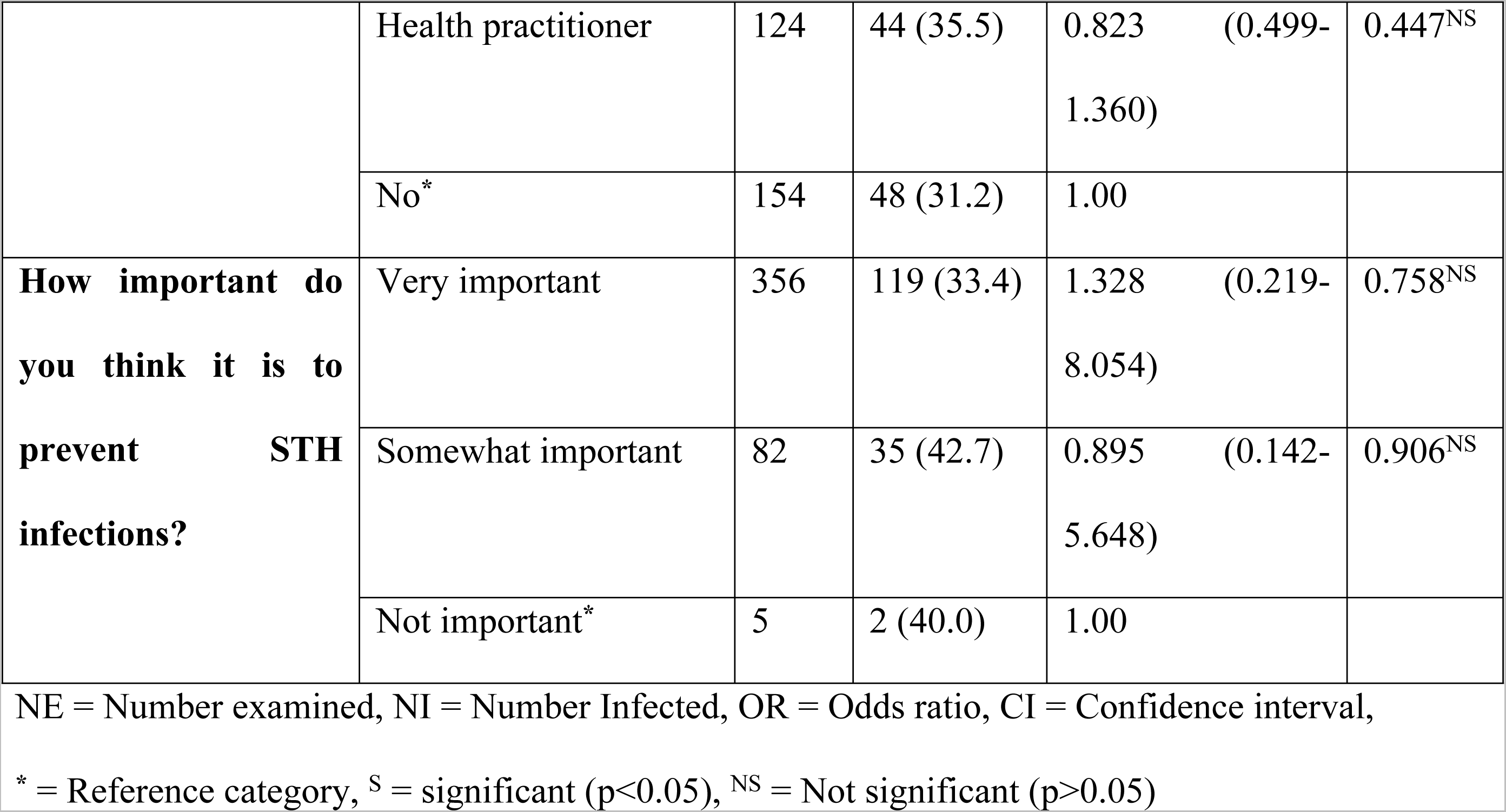
Multinomial logistic regression analysis of risk factors associated with any soil-transmitted helminths infection among adolescents studied in Anaocha L.G.A., Anambra State (n=443)

Adolescents who had no idea how STH infection was transmitted (OR: 5.526; 95% CI: 3.298-9.270; p<0.05) were 5.5 times more likely to be infected (Fig. 4) (Table 4).

**Fig. 4:**
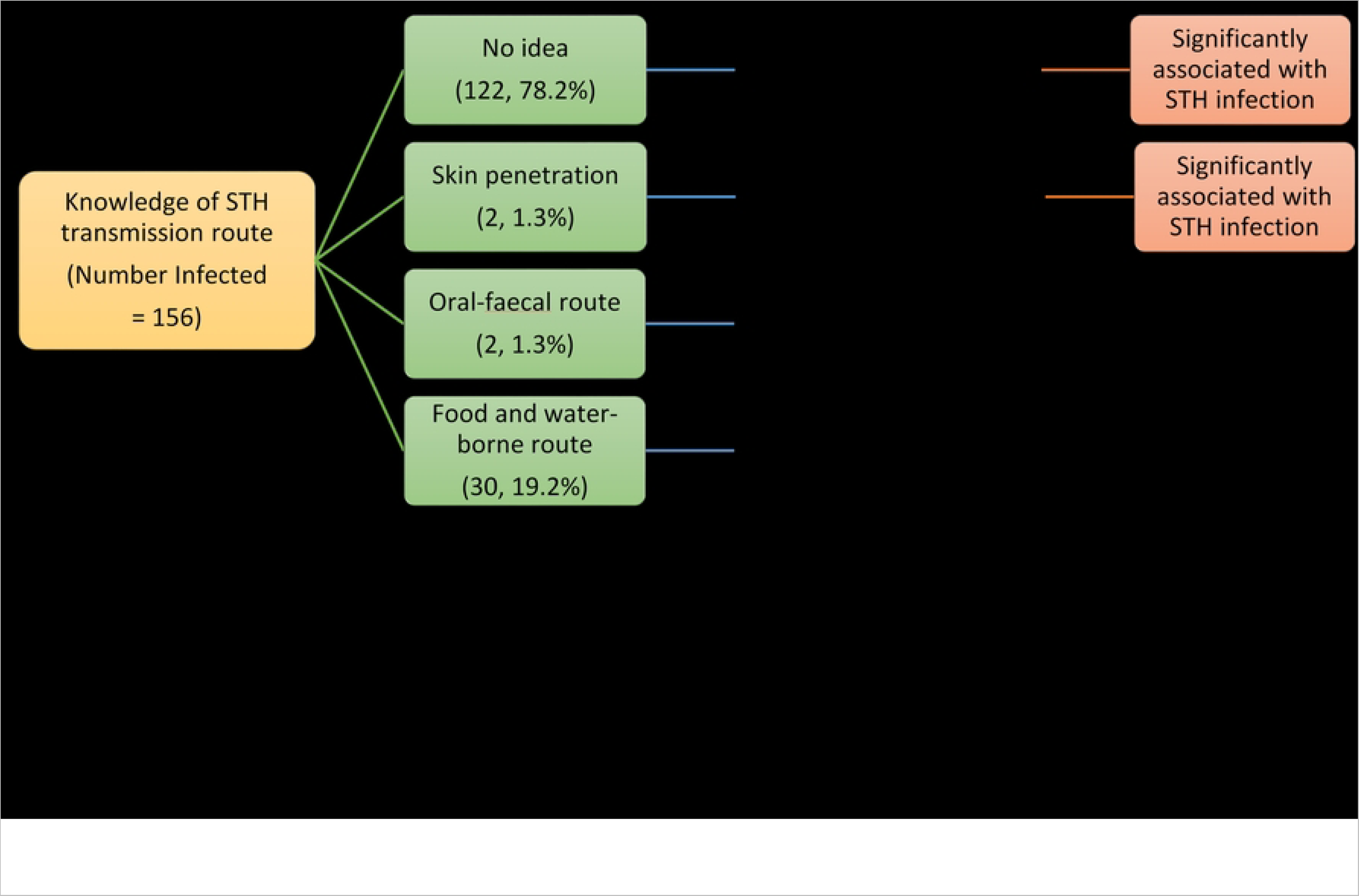
Association of knowledge of infection transmission route with soil-transmitted helminths infection among adolescents studied in Anaocha L.G.A., Anambra State.

Regarding STH infection with parents’ occupation indicates that adolescents whose parents were traders/entrepreneurs (OR: 7.856; 95% CI: 1.600-38.559; p<0.05) and teacher/civil servants (OR: 6.000; 95% CI: 1.140-31.590; p<0.05) were significantly more likely to be infected (Fig. 5) (Table 4). Another significant finding was the level of education of their parents, those whose parents completed only primary education (OR: 2.561; 95% CI: 1.274-5.150; p<0.05) were 2.6 times more likely to be infected with STH infection compared to those whose parents attained tertiary education (Fig. 6) (Table 4).

**Fig. 5:**
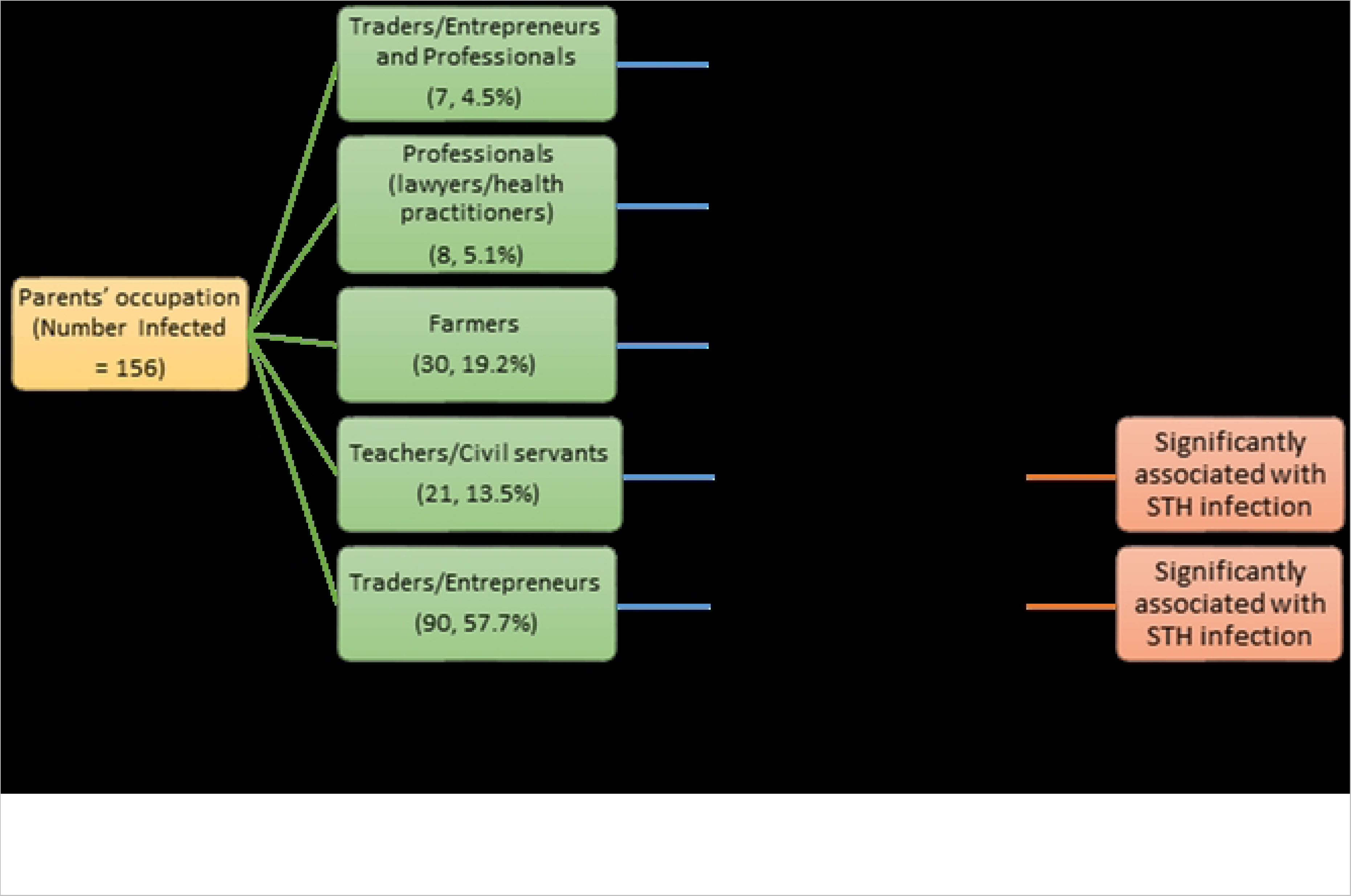
Association of parents’ occupation with soil-transmitted helminths infection among adolescents studied in Anaocha L.G.A., Anambra State.

**Fig. 6:**
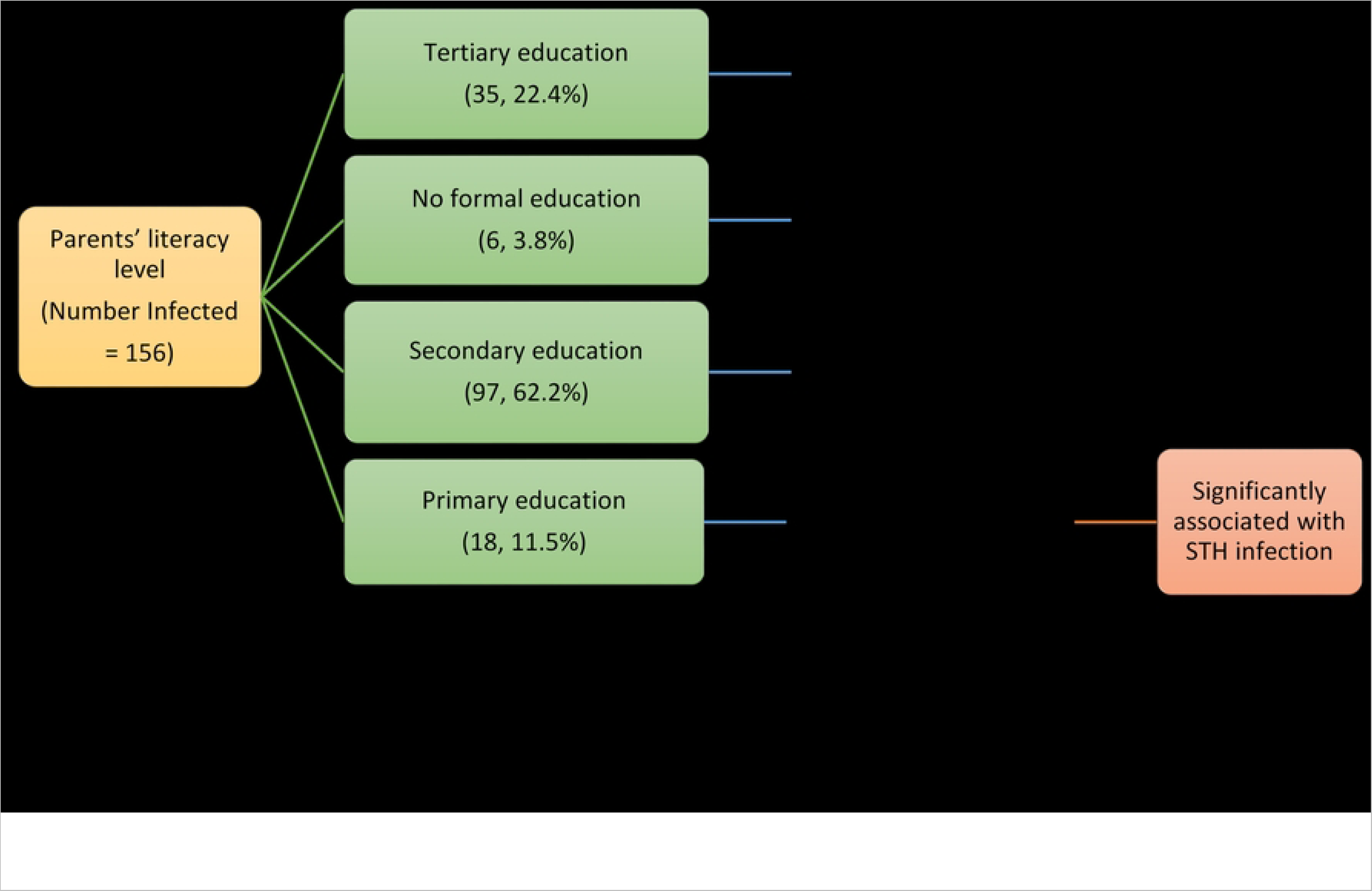
Association of parents’ literacy level with soil-transmitted helminths infection among adolescents studied in Anaocha L.G.A., Anambra State.

Adolescents in JSS 1 class (OR: 0.291; 95% CI: 0.152-0.556; p<0.05) and JSS 3 class (OR: 0.336; 95% CI: 0.148-0.764; p<0.05) were significantly less likely to be infected compared to adolescents in SSS 2 class (Fig. 7) (Table 4). Adolescents who used pit toilets (OR: 3.750; 95% CI: 1.542-9.119; p<0.05) and water cisterns (OR: 2.671; 95% CI: 1.159-6.158; p<0.05) were more likely to be infected than those who practised open defecation (Fig. 8) (Table 4).

**Fig. 7:**
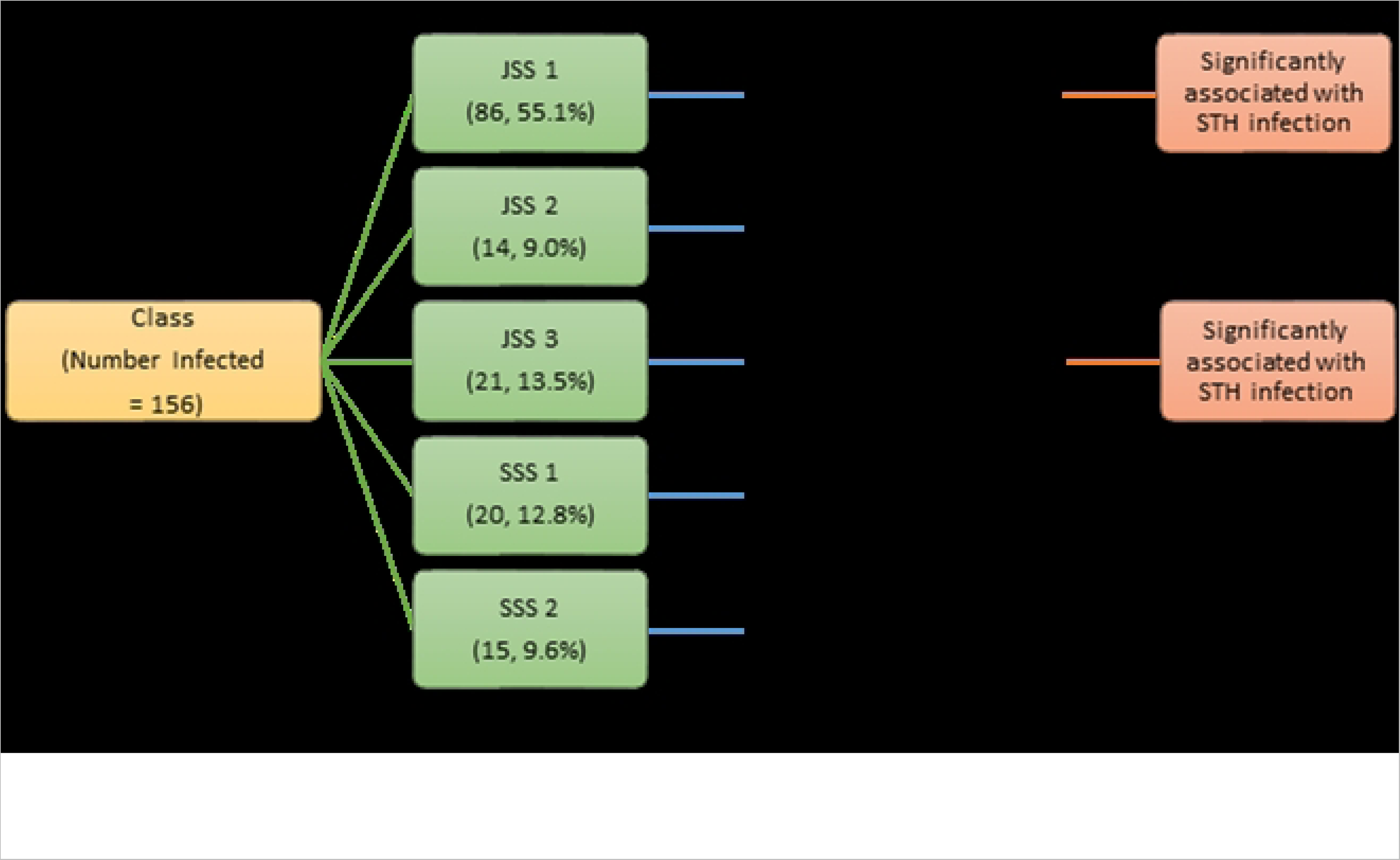
Association of adolescents’ class in school with soil-transmitted helminths infection among adolescents studied in Anaocha L.G.A., Anambra State.

**Fig. 8:**
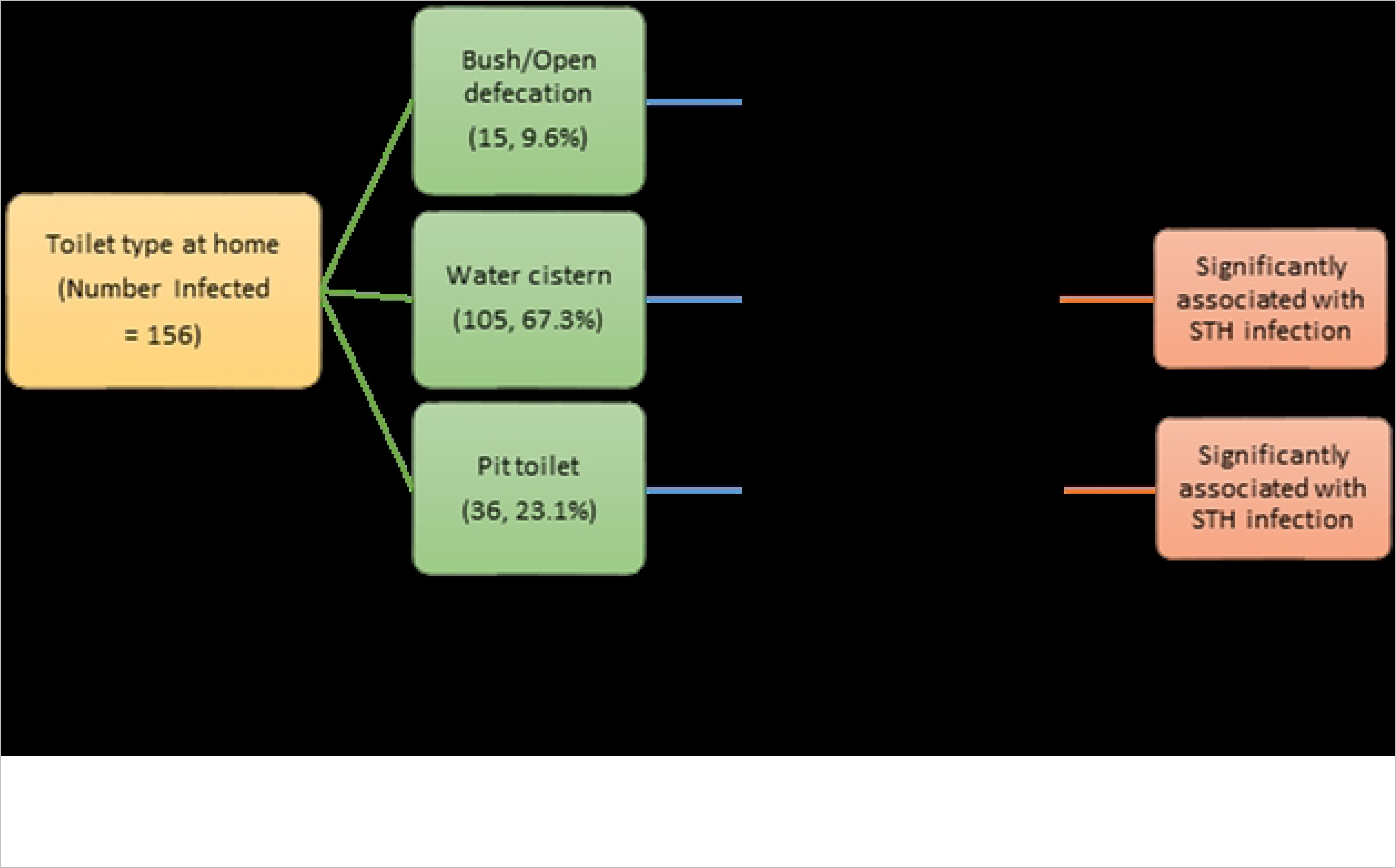
Association of type of toilet in the home with soil-transmitted helminth infection among adolescents studied in Anaocha L.G.A., Anambra State.

Gender, age, source of drinking water, biting nails, washing hands before and after meals, and washing fruits did not have any significant statistical relationship with positive diagnosis of STH infection.

## Discussion

This study found that the overall prevalence of soil-transmitted helminths (STH) among adolescents in Anaocha LGA was 35.2%, which classifies the area as a “moderate risk area” for preventive therapy based on WHO standards [21]. This suggests that Adazi-Nnukwu and Agulu are endemic for STH infections. Despite efforts to control STH infections through mass drug administration (MDA) since 2013 in Anambra state, the infections persist among adolescents. This may be due to potential gaps in school-based MDA, which primarily target nursery and primary school-aged children, resulting in many adolescents in secondary schools being overlooked. The selective approach may have contributed to the notable prevalence of STH among adolescents, posing challenges in completely eradicating this disease. Various studies also suggest that the targeted intervention approach hinders the control and elimination of infections [22–24]. This is because infected adolescents may act as significant carriers of this infection, contributing to its continuous transmission within the community.

The prevalence observed in this study was higher compared to previous research conducted in Anambra State [11, 13–14, 25–26]. However, it was lower than the prevalence reported in the studies conducted by [8] and [27] respectively, also within Anambra State. The dissimilarities could be attributed to variations in socio-cultural factors, behavioural traits, climatic conditions, the implementation of prevention and control measures, and the frequency and effectiveness of MAMs in the different communities. Additionally, variations in diagnostic techniques used in the studies may have also contributed to the differences observed.

The predominant helminth parasite was *Ascaris lumbricoides* (16.9%), followed by *Trichuris trichiura* (1.4%) and hookworm (0.5%). These findings align with earlier studies that also reported *A. lumbricoides* as the most prevalent STH [11–12, 25, 28–29] but contrasted with [26] and [30] who reported hookworm as the most prevalent STH. Thus, the high prevalence of *A. lumbricoides* among adolescents in this study highlights its continued dominance as the primary helminth parasite in Anambra state. This can be attributed to the long lifespan and high fecundity rate of the female worm, which results in the daily release of a large number of eggs into the environment. Furthermore, the embryonated eggs of *A. lumbricoides* are highly resilient to harsh environmental conditions compared to other STH.

Studies show that most cases of helminthic infections in Africa occur in the form of co-infections rather than isolated infections due to common epidemiological factors that facilitate the spread of these diseases [31–33]. In this study, 16.5% of adolescents studied had multiple infections, which was higher compared to previous studies conducted in southern Nigeria [12, 31, 34–35]. This is not surprising, especially in areas where helminth infections are endemic, as prolonged exposure increases the likelihood of acquiring multiple infections. Notably, the most predominant co-infection was *A. lumbricoides* and hookworm (10.8%) which is consistent with the findings of [30, 32]. Furthermore, 2.5% of the study population harboured all three STH species simultaneously. Common factors that could contribute to these co-infections are low standard of living and poor hygiene practices [32–33]. These findings are particularly important because individuals with multiple infections may be at a higher risk of experiencing multiple health problems and significant morbidity associated with helminth infections. A noteworthy incidental observation made during this study involved a female adolescent harbouring a co-infection of *A. lumbricoides* and hookworm, who also passed uric acid crystals in her stool and was found to have pubic lice (*Pthirus pubis*). These additional findings may indicate the presence of underlying health conditions that require further investigation.

All the adolescents who participated in the study had light-intensity infections of *A. lumbricoides*, *T. trichiura* and hookworm, which is consistent with previous reports [11–13, 31, 36]. The absence of heavy-intensity infection indicates a low transmission risk and could be attributed to the ongoing MAMs in the state. However, people with light-intensity infections often do not seek treatment due to the absence of noticeable symptoms. This could contribute to the contamination of the environment and the sustained transmission of the parasites in the community. Therefore, it is important to treat adolescents with light-intensity infections, even if they do not have any symptoms. This will help to reduce the burden of parasitic eggs and the prevalence of the disease. The intensity of hookworm and *A. lumbricoides* infections was observed to be higher in females, except for infection with *T. trichiura* which was higher in males. Conversely, the intensity of hookworm was higher in adolescents aged 10-14 years, contrary to *A. lumbricoides* and *T. trichiura* which were higher among those aged 15-19 years. However, none of these changes were statistically significant. More study is required to investigate the potential factors underlying the observed gender and age disparities in the severity of helminth infection among adolescents.

This study revealed no significant correlation between gender or age and STH infection. This indicates that the infection is not specific to any particular gender or age group and that adolescents have similar levels of exposure to predisposing factors. These findings are consistent with previous studies conducted in Nigeria [11, 13, 25, 37–39].

The study found that while STH infections were more prevalent among adolescents in JSS 1, they were the least likely to be infected with STH among the study population. This could be attributed to the previous deworming medicines they had received during MAMs programs while in primary school which conferred enhanced protection as they transitioned into secondary school. However, poor personal hygiene habits among those in JSS 2 may have contributed to the increased risk of infection observed in this group. In contrast, JSS 3 and SSS 1 students had lower chances of infection, possibly because they had become more aware of their hygiene and as senior students, were expected to maintain higher levels of cleanliness and hygiene. These findings highlight the significance of MAMs programs and the immediate need for improved hygiene practices to prevent the spread of STH infections.

The study identified potential risk factors for STH infection (Fig. 9). Adolescents who had no knowledge of STH had higher odds of being infected, than those who had some knowledge. Similar conclusions were reached by other studies, which also identified insufficient knowledge about STH infections as a factor associated with the occurrence of the infection [13, 31, 40–41]. This is because a lack of understanding about STH infections makes individuals more susceptible to infection since they are unaware of the factors that expose them to the infection. Therefore, it is crucial to incorporate health education and sensitization into existing intervention programs to increase awareness. Parental socioeconomic status has been identified as a crucial risk factor for STH infections in school-aged children [39]. This study observed a significant association between parents’ occupation and STH. Adolescents whose parents were traders or entrepreneurs were 7.9 times more likely to be infected with STH, while those whose parents were teachers and public servants were 6 times more likely to be infected. Previous studies however, have reported higher odds of infection among school children whose parents were farmers [28, 42–44]. Similarly, the likelihood of acquiring STH infection was higher among adolescents whose parents/guardians had attained primary education than those whose parents/guardians had no formal education, although the observed prevalence was lower in the primary group compared to the no formal education group. This is in contrast with the reports of [31, 45] but supports the argument of [40] who suggested that this may be due to parents spending more time on outdoor activities to provide for their families, resulting in less time spent with their children. As a result, adolescents may have poor personal and food hygiene and engage in outdoor activities that increase their risk of STH infection.

**Fig. 9:**
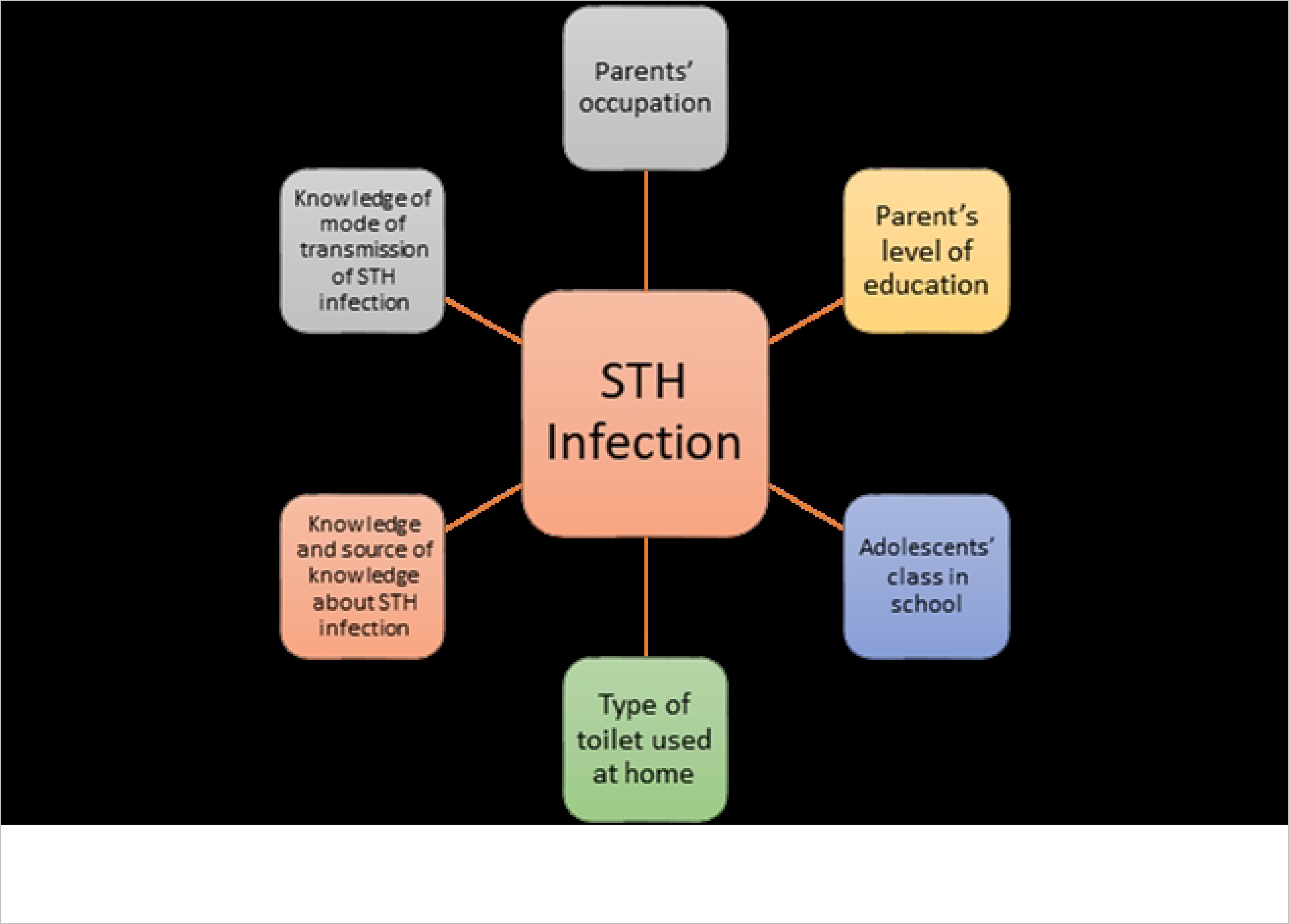
Risk factors associated with soil-transmitted helminths infection among adolescents studied in Anaocha L.G.A., Anambra State.

The likelihood of infection with soil-transmitted helminths in adolescents who did not have toilet and water facilities in school was higher than in adolescents who did. This was expected as the schools visited in the study area had no toilets available and substandard water facilities. This leads to indiscriminate defecation by adolescents in the school surroundings exposing them to infections with soil-transmitted helminths. This concurs with findings from similar studies conducted in Nigeria [12, 26–27, 31]. Using water cisterns and pit toilets at home was significantly associated with higher odds of infection, which contradicts the findings of other studies [28, 40, 46–48]. The higher infection odds observed for adolescents who use water cistern is not surprising since the majority of the adolescents who tested positive for STH infections do not wash their hands after defecation. This could be due to a lack of toilet and water facilities in the schools attended by the adolescents in the study, causing them to resort to open defecation in nearby bushes around their schools, thus fostering unsanitary habits during their school hours and might increase their risk of infection despite the use of water cisterns at home. Additionally, this could be linked to the improper maintenance and poor sanitation of the toilet. Although water cisterns are renowned for their protective attributes against STH, their effectiveness diminishes if it is not provided alongside adequate water supply to ensure personal cleanliness and cleanliness of the latrine, increasing the chances of faecal contamination as has been reported by [13].

It is interesting to note that adolescents who had received deworming treatment 1-3 months before the study had significantly lower odds of infection compared to those who had received treatment more than 6 months before the study underlining the pivotal role of regular deworming treatments to substantially reduce the risk of STH. However, there was a high prevalence of STH infections among adolescents who had been dewormed indicating cases of reinfection. This finding is consistent with a study conducted in rural Ghana [49] and may be attributed to potential resistance to the recommended medication and its efficacy. It is important to note that anthelmintic drugs have varying levels of effectiveness against different species of STH, and they are particularly ineffective against *Trichuris trichiura* when using single-dose treatments [50]. Previous comparative studies have shown that a triple-dose regimen is more effective in treating STH than a single or double-dose regimen, particularly in co-endemic areas where all three STH species are prevalent [51–53]. Additionally, a study found that the combination therapy of ivermectin-albendazole had good tolerability and higher efficacy against *T. trichiura* compared to the current standard single-dose albendazole treatment, supporting its use in PC programs [54].

## Conclusion

Over one-third of the adolescents studied in Anaocha Local Government area, Anambra State, were infected with at least one STH species. Although the intensity of STH was low, there is a need for a more comprehensive preventive approach that includes all adolescents in the study area. This is because adolescents could act as significant reservoirs for reinfection of treated children and other at-risk groups in the community, posing serious challenges to the target of eliminating STH as a public health issue in Anambra State. Therefore, it is imperative to revise the treatment strategy and prioritize the inclusion of adolescents in all MDA interventions in Anambra state, Nigeria. Additionally, implementing innovative intervention strategies aimed at controlling or preventing STH infection is crucial. Health education programs utilizing creative mediums such as theatre, art, and music, which involve adolescents as educators and advocates, should be developed to enhance community engagement. Alongside this, embracing innovative toilet designs, waste management systems, and sanitation technologies that are both cost-effective and sustainable at the community and household levels is essential. Furthermore, there is a need to initiate behaviour change campaigns that employ novel approaches like gamification, mobile apps, and social media. These platforms can effectively educate adolescents about the significance of hygiene practices, regular deworming, and proper sanitation. By harnessing these innovative techniques, a more comprehensive and impactful approach to combating STH infections and improving public health in Anambra State and similar states in the country can be attained.

## Limitations

Intervention programs should be conducted at a time when all demographic groups can participate to avoid exclusion as was the case in this study where the senior secondary class 3 (SS3) was not included in the study due to the ongoing West African Senior Secondary Examinations (WASSCE) at the time of the study. Additionally, the laboratory technique used in this study, the Kato-Katz technique, has limited sensitivity to light-intensity infections and cannot accurately detect *Strongyloides stercoralis*, which may have resulted in missing some infections. Furthermore, while this technique is acknowledged for its heightened sensitivity for *A. lumbricoides* and *T. trichiura*, it is less sensitive for hookworms, which may have contributed to the low prevalence of hookworm infection reported in this study.

## Data Availability

All data produced in the present work are contained in the manuscript. Relevant data that does not violate the rights of the participants has been attached as supplementary files. For any inquiries or requests regarding the data, please contact [Ogechukwu B. Aribodor] at

## Acknowledgements

We would like to extend our heartfelt appreciation to the Health Research and Ethical Committee at Chukwuemeka Odumegwu Ojukwu University Teaching Hospital (COOUTH), the Head of NTDs at Anambra Ministry of Health, the Chairperson and Head of Research, Post Primary Schools Service Commission (PPSC) for facilitation and support of this project. We are also thankful to the laboratory technologists of the Department of Zoology, Nnamdi Azikiwe University, Awka for collecting and examining the stool samples. We are grateful to the school principals, teachers and students of Community secondary school, Adazi-Nnukwu and Union secondary school, Agulu for their cooperation in participating and providing the necessary information and stool samples. Without them, this research project would not have been possible.

## Supporting Information

